# Biobank-scale survey of gene-diet interactions informs precision nutrition polygenic scores

**DOI:** 10.64898/2026.04.13.26350340

**Authors:** Matteo Di Scipio, Alice Man, Ricky Lali, Jianhan Wu, Ann Le, Paul W. Franks, Guillaume Paré

## Abstract

Genome-guided dietary advice is a goal of precision nutrition. However, the contribution of gene-diet interactions (G×Ds) to disease risk remains unclear, hindering the identification of diet-outcome pairs more likely amenable to genetic-based recommendations. We thus implemented a two-step approach: first, we comprehensively assessed the contributions of genome-wide G×Ds to cardiometabolic outcomes across a broad array of dietary exposures in UK Biobank participants (*N* = 141,144 to 325,989). Second, we selected the 20 significant diet-outcome pairs from the 713 pairs tested (*p* < 7.0 × 10^-5^) and derived G×D polygenic scores. In an independent sample, all scores were nominally associated with their corresponding outcomes, with 12 of 20 polygenic scores Bonferroni significant (p < 0.0025). Further analyses revealed G×D polygenic scores were associated with clinical outcomes such as incident gout, suggesting translational potential. Altogether, these results showcase the promise of G×D scores to inform precision nutrition.

## Introduction

An overarching goal of precision nutrition is to reduce the global burden of nutrition-related non-communicable diseases by providing individually tailored dietary recommendations^1^. Traditional dietary guidelines, which are based on population averages, often overlook meaningful inter-individual differences in response to dietary interventions^2^. Nutrigenomics studies suggest that genetic factors may contribute to these inter-individual differences^2,3^, in addition to other precision nutrition components such as microbiome profiling, behavioral and environmental data, digital health tools, and AI-driven modeling^1^. However, evidence to date indicates that the predictive utility of SNP-based models in real-world nutrition settings remains modest^3,4^. Nonetheless, there is growing interest in integrating genetic information into nutrition algorithms, and this could be in the form of gene–diet interaction (G×D) polygenic scores^5–7^. Despite their potential, G×D polygenic scores remain underutilized due to limited predictive performance and insufficient clinical validation.

Although there have been recent advancements in gene-environmental (G×E) scores^8,9^, developing and applying G×E PRS models faces several challenges. These include deriving polygenic interaction terms^10^, model misspecification^11^, multiple testing burden^12^, and the need for computationally efficient methods ^12^. In the context of G×D, a much less discussed but equally critical limitation is the lack of a comprehensive survey of trait-diet pairs with appreciable genome-wide G×D effects, which can help prioritize specific traits and dietary exposures for constructing G×D scores^13^. To address these challenges, we propose a two-step approach: first, to systematically evaluate the variance in outcomes explained by genome-wide G×D across a broad range of cardiometabolic outcomes and dietary exposures in biobank-scale data; and second, to construct and apply genome-wide G×D polygenic scores for trait-diet pairs with statistically significant genome-wide G×D effects. Genome-wide interactions studies^14–16^ and linear mixed models^13,17,18^ have investigated G×D, though are not without limitations nor did they substantially progress to study the utility of interaction scores. On the other hand, studies constructing G×D polygenic scores for traits like cancer, diabetes, and anthropometric outcomes have yielded modest associations^8,19,20^, which may be owing to small discovery and validation sample sizes and lack of systematic prioritization of diet-outcome pairs with strong G×D evidence. Additionally, these scores often fail to account for genome-wide interactions comprehensively, limiting their predictive power and applicability to precision nutrition. For instance, polygenic risk scores typically identify interactions only after selecting SNPs conveying statistically significant marginal associations, overlooking G×D that occur outside these marginal effects. This limits their ability to capture the full complexity of gene-diet interplay and likely contributes to their low predictive power^21^. In contrast, our approach systematically leverages G×D estimates to prioritize all SNPs with evidence of interactions.

In this report, we estimated genome-wide G×D with MonsterLM^21^, a G×E method for unbiased genome-wide interaction variance estimation, using a large-scale, unparametrized approach that improves statistical power while minimizing bias. We quantified genome-wide G×D across 713 trait-diet pairs comprising 31 clinically relevant traits and 23 dietary exposures, including single-nutrient intakes (SNIs), dietary indexes (DIs), and global dietary principal components (dPCs) from the UKB food frequency questionnaire. Filtering for significant trait-diet pairs, we constructed polygenic G×D scores and assessed their relevance for precision nutrition. This framework provides a robust foundation for identifying diet-outcome pairs with favorable precision nutrition translation potential.

## Results

### Baseline Dietary Characteristics in the UK Biobank

Our study comprehensively quantifies genome-wide G×D in the UKB. We tested 31 health and anthropometric outcomes for gene-diet interactions using 23 diet variables, selected for their coverage of major food groups and established relevance to cardiometabolic traits (**Figure 1**). After inclusion and exclusion criteria (**Figure 1**), up to 325,989 unrelated white British participants were used to calculate G×D for outcomes with dPCs, DIs and SNIs (derived from the 24-hour diet recall). In total, dietary exposures were subdivided into eight classes, including dPCs, DIs, and six SNI classes: carbohydrates, discretionary, fats, minerals, proteins, and vitamins. Participants were divided into discovery and validation sets, with 80% used for analyses (up to 260,791) including the G×D screen and G×D score derivation, and 20% (up to 65,198) used for testing the derived G×D scores (**Extended Data Figure 1**).

**Figure 1.**
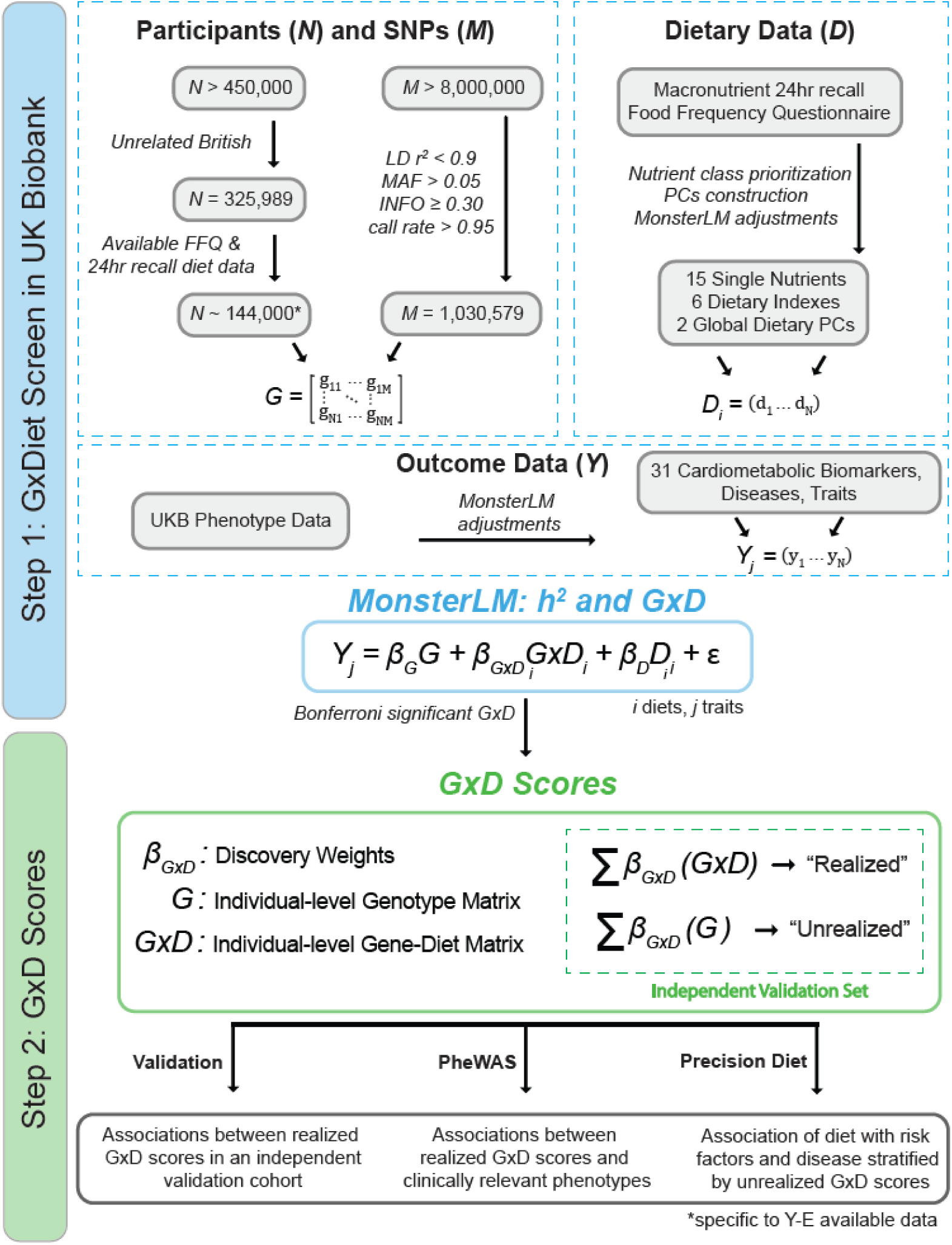
| Full study design illustrating all analyses workflow. The preliminary step (top panel) describes participant quality control and filters as well as dietary data sources and selection. The MonsterLM algorithm (blue panel step 1) was used to calculate heritability and G×D for each outcome-diet combination. Then significant findings were used to develop, validate, and apply G×D scores (green panel step 2). Significant Bonferroni G×D underwent follow-up analyses (bottom panel) including validation, PheWAS, and precision diet tests.

The baseline characteristics of all participants are shown in **Table 1**. When sorted by the Healthy Eating Index 2020 diet index, participants in the highest tertile were slightly older (mean 57.2 years of age versus 56.4 and 56.5 in lower tertiles) and more likely to be females (60.2%), whereas female representation in tertile 1 and 2 were 48.2% and 54.3%, respectively. Six DIs (Dietary Approaches to Stop Hypertension [DASH], Healthy Eating Index-2020 [HEI-2020], Alternative Healthy Eating Index [AHEI], Mediterranean Diet Index in serving sizes from the PREDIMED trial [MEDI], Dietary Inflammatory Index [DII], American Cancer Society 2020 dietary index [ACS 2020]) were calculated in the second cohort with more extensive nutrient intake data. When comparing HEI2020 tertile 3 (HEI-T3) to tertile 1 (HEI-T1) across the other five DIs, some trends were observed: compliance with DASH, AHEI, and MEDI diets was greater in HEI-T3 compared to HEI-T1; the HEI-T3 cohort’s diet was significantly more ‘anti-inflammatory’ (with a more negative score on DII) than that of HEI-T1; and the HEI-T3 cohort generally had higher ACS scores, aligning with a recommended diet for cancer risk reduction (**Table 1**). A correlation matrix between all DI scores shows considerable correlation between the DIs with the average pairwise correlation (Pearson’s *r*) between DI ranging from 0.265 to 0.508 (**Extended Data Table 1**). Correlations were highest for DASH, HEI2020, AHEI, and ACS scores, ranging from 0.532 to 0.737 (**Extended Data Table 1**).

**Table 1.** All Participants Baseline Characteristics, Dietary Indexes and Dietary Patterns.

|  | **Total (FFQ)** | **Total (DIs)** | **HEI Tertile 1** | **HEI Tertile 2** | **HEI Tertile 3** |
| --- | --- | --- | --- | --- | --- |
| *N* | 325,989 | 141,144 | 47,048 | 47,048 | 47,048 |
| Age (years); range | 56.86; 39 – 72 | 56.4 (7.9); 39 – 72 | 55.4 (8.1); 39 – 70 | 56.5 (7.8); 40 – 72 | 57.2 (7.6); 40 – 70 |
| Sex, *N* |  |  |  |  |  |
| men | 150,741 | 64,597 (45.8) | 24,381 (51.8) | 21,496 (45.7) | 18,721 (39.8) |
| women | 175,248 | 76,547 (54.2) | 22,667 (48.2) | 25,552 (54.3) | 28,327 (60.2) |
| **Dietary Indexes** |  |  |  |  |  |
| DASH | - | 24.5 (3.7); 12 – 36 | 22.4 (3.2); 12 – 34 | 24.5 (3.1); 12 – 36 | 26.7 (3.2); 13 – 36 |
| HEI2020 | - | 47.3 (11.1); 0 – 87.8 | 35.3 (5.4); 8.7 – 42.2 | 47.1 (2.8); 42.2 – 52.0 | 59.6 (5.9); 52.0 – 87.8 |
| AHEI | - | 60.2 (9.4); 20.8 – 95.8 | 55.6 (8.4); 20.8 – 88.4 | 60.1 (8.6); 24.3 – 93.9 | 64.8 (8.7); 31.3 – 95.8 |
| MEDI | - | 3.80 (1.4); 0 – 10 | 3.16 (1.1); 0 – 8 | 3.74 (1.22); 0 – 9 | 4.5 (1.39); 1 – 10 |
| DII | - | -1.53 (0.8); -4.2 – 4.8 | -1.29 (0.8); -3.8 – 4.8 | -1.59 (0.8); -4.2 – 4.6 | -1.72 (0.8); -4.1 – 2.0 |
| ACS | - | 6.02 (2.3); 0 – 12 | 4.44 (1.9); 0 – 11.8 | 5.98 (1.9); 0 – 11.8 | 7.64 (1.9); 0.3 – 12 |

**Cohort Descriptive Characteristics.** Genome-wide G×D estimates were obtained across 141,144 unrelated British individuals from the UKB. Descriptions of the cohort are presented as means with standard deviation and range. A subset of the total cohort is presented in tertiles ordered by their HEI2020 scores. All descriptive statistics are from baseline data except from the 24-hour diet questionnaire. *N*: Number of Individuals. DASH: Dietary Approaches to Stop Hypertension; HEI2020: Healthy Eating Index 2020; AHEI: Alternative Healthy Eating Index; MEDI: MED Index in serving sizes from the PREDIMED trial; DII: Dietary Inflammatory Index; ACS: American Cancer Society.

Dietary patterns were also quantified by calculating FFQ-derived principal components (dPCs) using 17 FFQ features (**Extended Data Table 2**). The total variance explained by the first 5 dPCs (out of 17) was 43.7%, with dPCs 1 through 5 explaining 13.5%, 9.9%, 7.2%, 6.6%, and 6.5% respectively. Feature loadings in dPC1 were largely weighted by beef, lamb/mutton, pork, and processed meat intake. dPC2 feature loadings include oily fish, non-oily fish, fresh fruit, and cooked vegetable intake.

### Genome-wide G×D Screen in the UK Biobank

We applied MonsterLM to estimate the genome-wide G×D variance between 23 dietary exposures and 31 health outcomes in the discovery set. Health outcomes included anthropometric traits and cardiometabolic blood biomarkers broadly related to glucose metabolism, inflammation, kidney disease, dyslipidemias, liver disease, serum electrolytes, sex hormones, and vitamins. All models were adjusted for age, sex, genetic principal components, and diet covariates used in the interaction term.

Of the 23 dietary exposures, we observed significant genome-wide G×D involving 4 dietary exposures, explaining between 2.1% and 21.2% of the total phenotypic variance in their corresponding health outcomes and belonging to 3 diet exposure classes: dPCs, DIs, and discretionary SNIs (**Figure 2**; **Extended Data Table 3**). Significant interaction estimates were found for BMI and urea in the G×dPC1 analysis. However, no significant estimates were observed for G×dPC2. Notably, apolipoprotein B showed a significant gene-diet interaction with the DI MEDI, representing the only significant G×D identified with DIs (**Figure 2; Extended Data Table 3**). Consistent G×MEDI results were observed in sensitivity analyses after accounting for lipid-lowering medication usage by exclusion of relevant participants or adjusting apolipoprotein-B levels (**Extended Data Table 4**). Two discretionary diet exposures tested, alcohol and caffeine (coffee), displayed 13 and 4 significant G×D estimates out of the 31 outcomes tested, respectively (**Figure 2**; **Extended Data Table 3**). The three most statistically significant GxAlcohol outcomes were apolipoprotein A, HDL cholesterol, and BMI with R^2^ estimates at 0.21, 0.21, and 0.17, respectively (**Extended Data Table 3**). The three most significant GxCaffeine outcomes were cystatin C, apolipoprotein A, and GGT with R^2^ estimates at 0.08, 0.07, and 0.05, respectively (**Extended Data Table 3**). No statistically significant G×D were observed for the other 19 dietary exposures. The outcome variance explained by the dietary exposures alone was also estimated and ranged from 0.00 to 0.051(**Extended Data Figure 2**).

**Figure 2.**
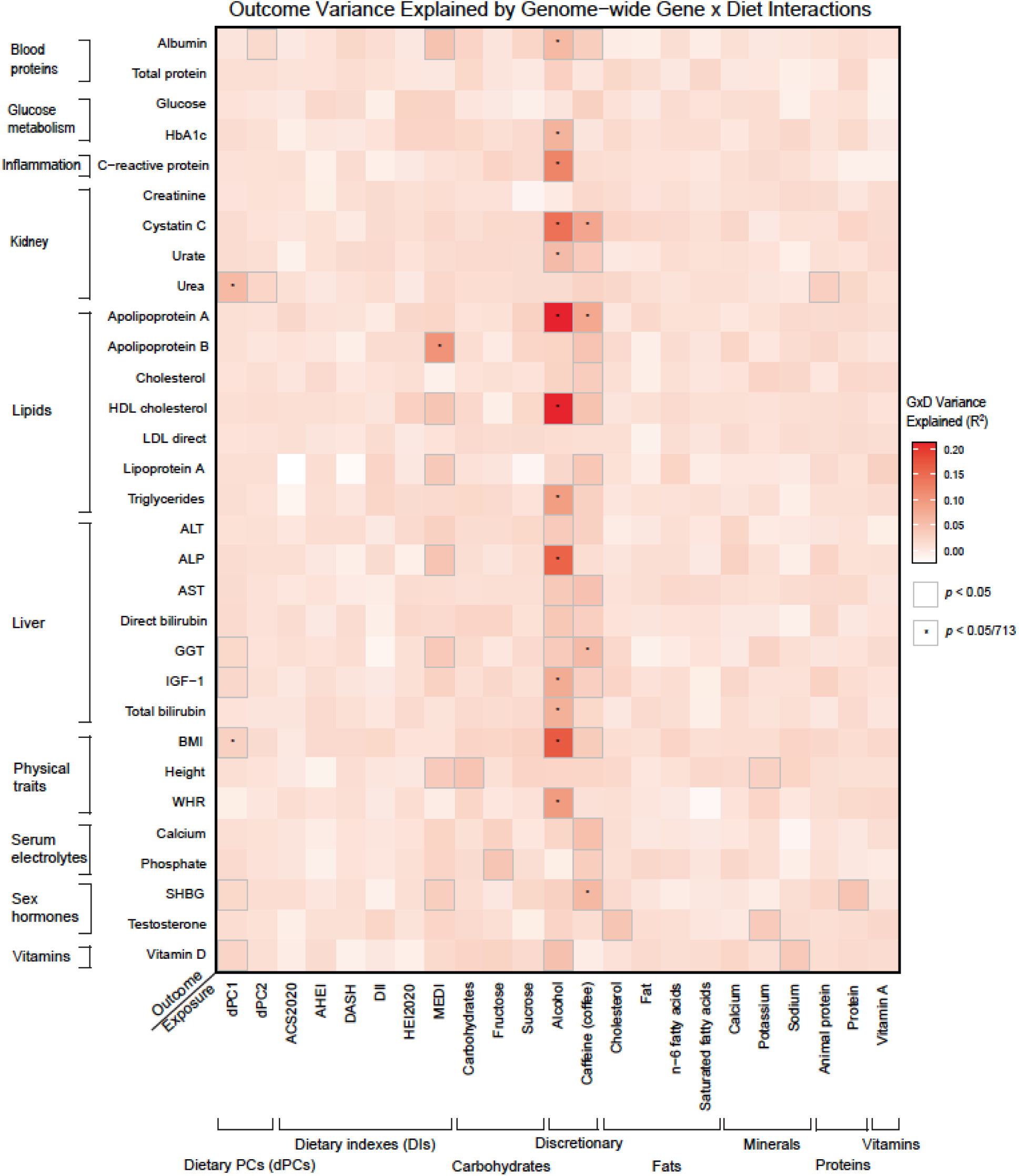
| Estimates of G×D across 23 dietary exposures and 31 health outcomes. Estimates computed using the MonsterLM G×E methodology. Outcomes are on the y-axis and dietary exposures are on the x-axis. Each cell contains the total G×E R^2^ estimate. Cells outlined in grey are nominally significant (*p*<0.05) and cells that are marked with an asterisk are Bonferroni significant (*p*<0.05/713).

Heritability (*h*^2^) of each 23 dietary exposures (dPCs, DIs, and SNIs) was estimated with MonsterLM. All dietary exposures were found to be significantly heritable, with *h*^2^ ranging from 0.038 to 0.088 (**Extended Data Table 5**). Heritability estimates for all 31 outcomes were significant and largely consistent with published estimates such as estimates from GRE^22^, SumHer^23^, and the LD Hub (LDSC) database^24^. As MonsterLM adjusts the outcome for any variance explained by the dietary exposure for each G×D analysis, heritability estimates for each outcome are shown for all exposure combinations and are virtually identical across all outcome-diet pairs tested (**Extended Data Figure 3**). The magnitude of GxE estimates are typically an order or more less than the variance explained by additive genetics (narrow-sense heritability)^21,25,26^. For the 20 G×D significant outcome-diet pairs tested, the ratio of G×D to *h*^2^ ranged from 0.16 to 0.70 (**Extended Data Figure 4**). Amongst the significant G×D pairs, many of them had alcohol as an environmental exposure. Therefore, alcoholic beverage subtypes were used as environmental exposures including beer/cider, spirits, and total wine (red and white). Beer/cider and wine-based drinkers consumed more than spirits across the cohort. Much of the larger magnitudes of G×D estimates include beer/cider and total wine as the exposures with spirits still showing significant but smaller magnitude estimates (**Extended Data Figure 5**).

### G×D Polygenic Scores and Diet-Outcome Associations

From the 20 significant G×Ds identified, we developed G×D polygenic scores to predict the corresponding outcome based on baseline diet exposure. G×D polygenic scores (thereafter G×D scores) were created by selecting SNPs from the MonsterLM analysis based on their interaction *p*-values in the G×D discovery set and weighting each SNP by their corresponding interaction regression coefficient under a clumping and thresholding framework (*C*+*T*)^27^. To avoid overfitting, G×D scores were tested in the validation set (independent from the discovery set) (**Figure 1; Extended Data Figure 1**). For each outcome-diet pair, five G×D scores were generated including SNPs with interaction *p*<0.25, *p*<0.1, *p*<0.05, *p*<0.01, and *p*<0.001, respectively. G×D scores for all 20 significant outcome-diet G×D results from the discovery cohort were computed, along with a null outcome-diet G×D serving as a negative control.

We tested the validity of the G×D polygenic score constructs in two different ways. First, for each participant, we multiplied the individual G×D score by the corresponding diet exposure to create a “realized G×D score”. For clarity, the G×D score (i.e. without multiplication with the corresponding diet exposure) is henceforth termed the “unrealized G×D score” (**Figure 1**). The realized G×D scores were associated with their corresponding outcomes with 12 out of 20 significant after adjustment for multiple hypothesis testing (*p* < 2.5 × 10^-3^) (**Figure 3**; **Extended Data Figure 6**). The strongest associations between the realized G×D score and outcome were at the interaction *p*-value threshold of 0.001, and henceforth this threshold was used in subsequent analyses (**Extended Data Figure 6**). For each major dietary class (SNIs, DIs, and dPCs) there was at least one significant association. For instance, amongst SNIs, a 0.13 SD unit increase in apolipoprotein A levels was observed per SD unit increase in the realized G×D score with alcohol as the dietary exposure (95% CI: 0.08 to 0.18, p < 0.001; **Figure 3**). In another example with caffeine as the interacting variable, the realized G×D score was associated with a 0.08 SD unit increase in cystatin C for each SD unit increase in the realized G×D score (95% CI: 0.02 to 0.14, *p* = 0.005; **Figure 3**). An SD unit increase in the realized G×D score for MEDI, a diet index, was associated with a 0.02 SD unit increase in apolipoprotein B levels (95% CI: 0.00 to 0.04, p = 0.042; **Figure 3**). Lastly, for dPC1, a dietary pattern PC, each SD unit increase in the realized G×D score was linked to a 0.07 SD unit rise in urea levels (95% CI: 0.02 to 0.13, p = 0.008; **Figure 3**). As a negative control, we also tested an outcome-diet combination that was non-significant in the main analysis, cholesterol-alcohol, and no association was observed. Further, unrealized G×D scores were not directly associated with either the diet or outcome on their own (i.e., univariate analysis) (**Extended Data Table 6**).

**Figure 3.**
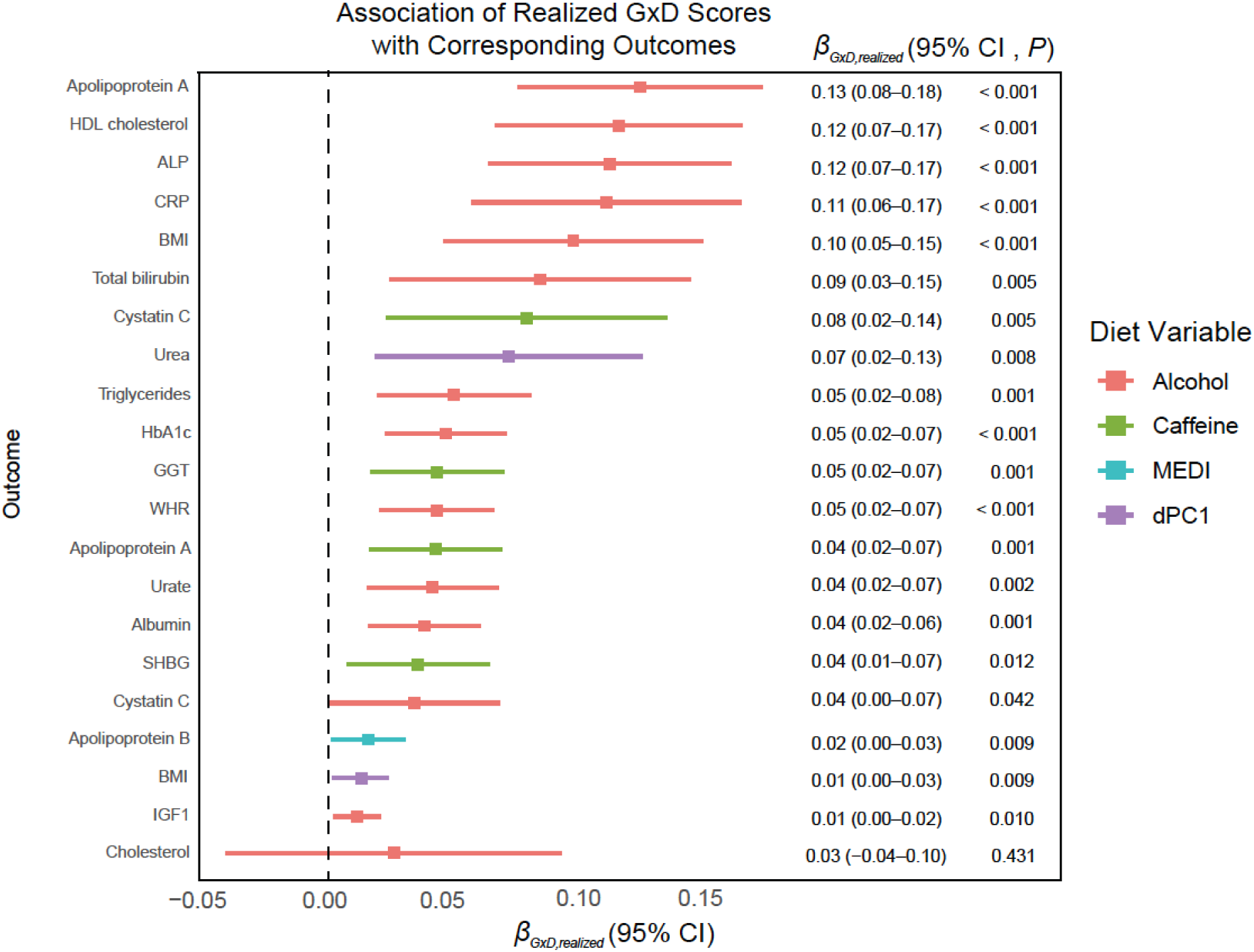
| Realized G×D Scores and Precision Diet Estimates. Estimates ( ) represent the regression coefficient from the validation cohort realized G×D score regressed onto the cohort’s outcome value. The point estimates represent the regression effect size of the G×D score regression and error bars denote 95% confidence intervals. All values are tabulated and shown to the right of each plot. The legend colours denote the diet environmental exposure. G×D scores were calculated with the *C*+*T* threshold method at discovery set selection of *p*<0.001. Twenty significant G D combinations were chosen from the initial screen, as well as one negative control, Cholesterol with Alcohol. Two levels of significance: nominally significant (*p*<0.05) and as Bonferroni significant (*p*<0.05/20) are considered. Validation cohort size was 20% that of the sample size used in the main screen (N∼28,000 per combination). G×D scores were generated using the *p*-value clumping plus thresholding approach (*P*_C+T_).

To test whether individuals can be stratified based on genetics associated with dietary effects, we examined how associations between dietary exposures and outcomes differed across deciles of unrealized G×D scores. Outcome–diet associations varied significantly across unrealized G×D score deciles for 8 of 20 combinations (p < 2.5 × 10^-3^) and showed nominal evidence of variation in 18 of 20 combinations (p < 0.05) (**Extended Data Figure 7**).

### Phenome-wide Association Study with realized G×D Scores

To further explore the broader implications of realized G×D scores, we performed a phenome-wide association study (PheWAS) in the validation set using the five most significant realized G×D from the preceding analysis (**Figure 3**). All realized G×D scores included alcohol consumption as the dietary exposure and the biomarker outcomes comprised BMI, alkaline phosphatase, apolipoprotein A, C-reactive protein, and HDL cholesterol. These five scores were then applied to a PheWAS analysis with over 300 PheCodes constructed with ICD-10 codes with a case count of at least 100, to assess their broader clinical relevance. Ten PheCodes were significantly associated with a realized G×D score after Bonferroni correction (**Table 2**). Importantly, the unrealized G×D scores without alcohol consumption were not associated with the corresponding PheWAS outcomes, highlighting the necessity of the interaction component (**Extended Data Table 7**).

**Table 2.** PheWAS of realized G×D scores (equation 5; alcohol diet variable) with ICD-10 phecodes.

| Phenotype | Case/Control Count | Odds Ratio (OR) | P-value | Outcome | 95% Confidence Intervals |
| --- | --- | --- | --- | --- | --- |
| Gout and other crystal arthropathies | 227/21532 | 1.317 | $5.60 \times 10^{-5}$ | BMI | 1.152, 1.506 |
| Abdominal hernia | 2647/19107 | 1.220 | $5.40 \times 10^{-5}$ | Alkaline phosphatase | 1.108, 1.343 |
| Gastritis and duodenitis | 1602/19942 | 1.309 | $1.10 \times 10^{-5}$ | Alkaline phosphatase | 1.161, 1.476 |
| Diaphragmatic hernia | 1503/19107 | 1.306 | $3.00 \times 10^{-5}$ | Alkaline phosphatase | 1.152, 1.481 |
| Other symptoms or disorders of the urinary system | 1236/18550 | 1.930 | $2.00 \times 10^{-5}$ | Apolipoprotein A | 1.586, 2.348 |
| Coronary atherosclerosis | 1048/18161 | 1.647 | $1.80 \times 10^{-5}$ | Apolipoprotein A | 1.354, 2.004 |
| Other chronic ischemic heart disease unspecified | 1312/18161 | 1.046 | $5.20 \times 10^{-5}$ | Apolipoprotein A | 0.86, 1.272 |
| Alcoholism | 489/20163 | 1.815 | $2.10 \times 10^{-6}$ | C reactive protein | 1.492, 2.208 |
| Type 2 diabetes | 967/18886 | 1.22 | $2.20 \times 10^{-5}$ | HDL cholesterol | 1.110, 1.420 |
| Diabetes mellitus | 1031/18886 | 1.20 | $5.40 \times 10^{-5}$ | HDL cholesterol | 1.097, 1.380 |

**Realized G×D PheWAS Estimates with Alcohol as the Diet:** The association between various ICD-10 phenotypes and the realized G×D score (with denoted outcome and exposures) as the independent variable. The *p*-values were adjusted for multiple comparisons (Bonferroni) depending on the phecode count used in each pheWAS. Realized G×D scores of $p < 0.001$ threshold was used.

Of the 10 associations identified, some involve highly prevalent chronic diseases. For example, the realized BMI – alcohol G×D score was significantly associated with gout and other crystal arthropathies (OR = 1.32 per SD, 95% CI: 1.152 to 1.506, *p* = 5.60 × 10^-5^) (**Table 2**), adjusted for BMI and alcohol. Similarly, coronary atherosclerosis was significantly associated with the realized apolipoprotein A – alcohol G×D score (OR = 1.65 per SD, 95% CI: 1.354 to 2.004, p = 1.80 × 10^-5^), adjusted for apolipoprotein A and alcohol. (**Table 2**). As a last example, the realized G×D score HDL cholesterol – alcohol was significantly associated with Type 2 diabetes (OR = 1.22, 95% CI: 1.11 to 1.42, *p* = 5.40 × 10^-5^), adjusted for HDL cholesterol and alcohol (**Table 2**).

### Gout as an example of a G×D score application

Gout affects over 55 million people globally and is a leading cause of inflammatory arthritis, characterized by debilitating flares and increased risk of cardiovascular and renal complications^28^. Among the associated conditions identified, gout stands out as a particularly compelling candidate for precision nutrition due to the strength of the BMI-alcohol G×D score association and alcohol’s known etiological link to gout^28^. Specifically, we ask: can the risk of gout associated with alcohol consumption be stratified using unrealized G×D scores?

To assess whether individuals with higher unrealized G×D scores experience differential effects of alcohol use on gout risk, we estimated the odds of incident gout across five quintiles of unrealized BMI-alcohol G×D score (**Figure 4a**). In individuals in the highest quintile of unrealized G×D score, each additional UK standard drink per day (18g EtOH) was associated with a 23.9% increase in odds of gout (95% CI: 1.11 to 1.38), while in the lowest quintile, the corresponding association was 4.8% (95% CI: 0.91 to 1.18). A significant trend was observed across the quintiles (p < 0.05) (**Figure 4a**). The model was adjusted for established gout risk factors, including age, sex, BMI, serum uric acid, hypertension, and serum creatinine.

**Figure 4.**
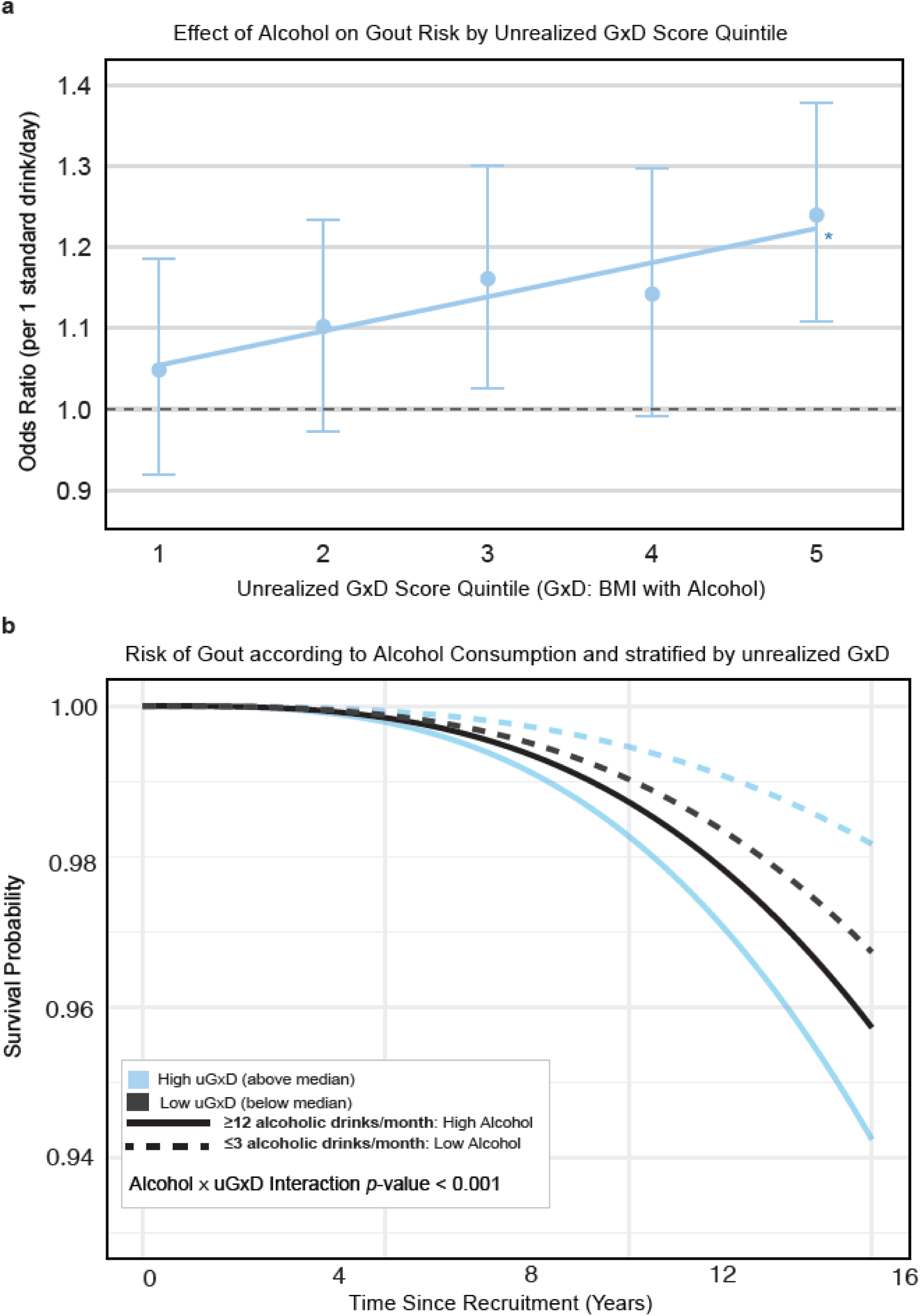
| Unrealized G×D Scores Stratify Incident Gout Risk in Relation to Alcohol Intake. a, Odds ratios (per 1 standard drink/day) for gout are shown across quintiles of the unrealized G×D score for BMI with alcohol. Point estimates reflect the per-drink effect size of alcohol on gout within each unrealized G×D score quintile. Vertical bars represent 95% confidence intervals. A significant positive trend was observed across quintiles (*p*<0.05*), with individuals in the highest quintile showing a 23.9% increase in gout risk per additional drink per day (p<0.001). **b,** Cox proportional hazards model of incident gout stratified by alcohol intake (High: ≥3×/week; Low: ≤3×/month) and unrealized G×D score (above vs below median). Curves are plotted over 15 years of follow-up. A global log-rank test confirmed significant differences across strata (p = 1.0 × 10ℒ³), and a significant interaction was observed between alcohol intake and G×D score group (p<0.001). 95% confidence intervals not displayed for ease of visualization. One alcoholic drink in the UKB FFQ approximates 18 grams of ethanol. uG×D: unrealized G×D.

To further evaluate the predictiveness of the unrealized G×D polygenic score for gout, we conducted a time-to-event analysis using a Cox proportional hazards model. Participants were stratified into four groups based on alcohol intake. “High” intake was defined as drinking ≥12 times per month (≥216 g ethanol/month, equivalent to ∼12 pints of beer), “Low” intake as ≤3 times per month (≤54 g ethanol/month) and into one of two unrealized G×D score bins (above vs below median score). Individuals in the “High Alcohol Intake” and “High unrealized G×D” group experienced the greatest risk of gout over the 15-year follow-up period (**Figure 4b**). As well, individuals with high unrealized G×D scores had substantially lower gout risk when alcohol consumption was low, suggesting that they may derive greater benefit from alcohol reduction. The interaction between alcohol intake and unrealized G×D group was significant (*p* < 0.001; **Figure 4b**).

### G×Alcohol Interactions Sensitivity Analyses

As alcohol yielded the greatest number of significant G×D combinations, we performed further analyses on the three most significant outcomes: apolipoprotein A, HDL cholesterol, and BMI. First, alcohol levels (in grams from 24-hour recall questionnaire) were randomly permuted in individuals who had any level of alcohol in their diet, while non-drinkers (no alcohol consumed reported) remained unmodified. As expected, the G×D estimates were attenuated (*p* = 1.4 × 10^-7^ to 3.4× 10^-15^) relative to no permutation (*p* = 2.5 × 10^-46^ to *p* = 1.0 × 10^-63^) for all three outcomes (**Extended Data Table 8**). When the alcohol intake value was fully permutated across all individuals, the G×D estimates were abrogated in all outcomes (*p* = 0.59 to *p* = 0.90). Null results were also obtained when randomly generating alcohol intake (**Extended Data Table 8**). Further, when alcohol consumption was treated as a binary variable (current drinker yes/no), G×D estimates remained significant and similar in magnitude to estimates recording alcohol consumption as a continuous trait (**Extended Data Table 8**). Lastly, to account for any potential confounding from socioeconomic status, we applied the Townsend Deprivation Index as an outcome covariate and the three G×Alcohol estimates remained significant (**Extended Data Table 8**).

Next, we tested whether the observed G×Alcohol effects were primarily driven by SNPs showing marginal associations with either the outcome or the alcohol exposure. Using UKB-independent GWAS studies, we extracted SNPs associated with alcohol^29^, HDL cholesterol^30^, and BMI^31^ according to five significance levels: *p* < 5 × 10^-8^, *p* < 1 × 10^-7^, *p* < 5 × 10^-2^, *p* < 1 × 10^-1^, and *p* < 1. Both heritability and GxAlcohol were estimated for the outcomes (apolipoprotein A, HDL cholesterol and BMI) at each of the 5 significance levels. Using the *p* < 1 set as the reference, the proportion of heritability and G×D variance recovered was more pronounced for heritability as compared to G×D, suggesting most G×D associations involve SNPs with weaker marginal associations with alcohol, HDL cholesterol, and BMI (**Extended Data Table 9**). For example, when testing BMI as the outcome, SNPs with marginal association *p*-values of *p* < 5 × 10^-8^, *p* < 1 × 10^-7^, *p* < 5 × 10^-2^, *p* < 1 × 10^-1^, and *p* < 1 with BMI recovered proportionally more heritability than for G×D (6.13%, 6.38%, 44.62%, and 55.34% versus 0.70%, 0.80%, 5.86%, and 8.51%, respectively) (**Extended Data Table 9**). Filtering SNPs for marginal association with the dietary exposure, alcohol consumption, serves as a test for potential collider bias. Collider bias occurs when a variable that is influenced by both the exposure and outcome is conditioned on, distorting the true relationship between the dietary exposure and outcome. As limiting the analysis to SNPs strongly associated with alcohol only modestly recovered G×D (0.70% to 8.51%), collider bias can be excluded as a sizeable source of spurious G×D as most of the interaction variance comes from SNPs without a strong marginal association.

## Discussion

A key goal of nutrigenomics is to develop tailored dietary recommendations. To do so, it is advantageous to first identify which diet-outcome pairs are subject to strong G×D effects as a foundation for developing predictive scores. Equally as important is identifying null G×D results in well-powered studies, because they highlight where *not* to look for genetically guided dietary recommendations. A major strength of our study is that it provides a broad assessment of genome-wide G×D through systematic testing of health-related outcomes with dietary exposures. Our results suggest a specific pattern of genetic architecture for G×D, with some evolutionarily recent dietary features (i.e. caffeine, alcohol) showing the strongest evidence for G×D. In turn, this points to trait outcome-diet pairs most likely to yield useful G×D polygenic scores.

Through our comprehensive survey of G×D comprising 23 dietary exposures and 31 outcomes in the UKB we identified significant G×D with dPC1, MEDI, alcohol, and caffeine, ranging from 2.1 to 21.3% of outcome variance. Notably, only 20 of 713 combinations tested yielded significant G×D, representing 2.8% of all combinations tested and suggesting that the presence of G×D is sparse. Hence, identifying *which* combinations have strong G×D is a critical, yet often overlooked, step into developing G×D scores. Our UKB G×D screen findings thus directed us *where* to develop G×D scores that hold potential for predicting dietary responses. Of the 20 G×D identified, we constructed G×D scores using clumping and thresholding (*C*+*T*) at various *p*-value cutoffs, identifying significant associations across all outcome-diet combinations in an independent validation set.

Our comprehensive G×D screen adds to the emerging literature surrounding G×D in biobank-scale datasets. For instance, G×Esum^25,32^ has been applied to search for genome-wide gene-diet interactions with 12 cardiometabolic outcomes and 20 dietary exposures (macronutrients, micronutrients, and food groups) and found negligible G×D contributions. Another study^17^ applied MTG2 on over 280,000 participants from the UKB with 12 diabetes-related metabolic traits and 8 lifestyle covariates identifying 12 significant G×E signals including two G×D combinations: BMI-alcohol and WHR-alcohol (also replicated by our study)^8^. By comparison, MonsterLM tested over 700 outcome-diet pairs—more than five times the combinations tested in these studies—leading to a greater number of significant signals. Additionally, MonsterLM captures individual-level genome-wide interaction variance without pre-filtering for marginal SNP associations, reducing bias and improving statistical power. Unlike summary-statistic and parametrized methods, which may underappreciate weaker or non-additive effects, our approach is more sensitive to detecting meaningful yet subtle G×D signals that previous studies may have missed^33^.

Subsequently, we developed G×D polygenic scores using a genome-wide, interaction-only SNP selection approach guided by G×D heritability. Unlike previous approaches that rely on marginal genetic effects typically from summary level GWAS data, our scores exclusively capture interaction-specific genetic variance from individual-level data^19,20,27,34^. Using MonsterLM-based clumping and thresholding (*C*+*T*) with a univariate *p*-value selection strategy, we prioritized SNPs with significant G×D. Other G×D scores studies have not found many appreciable signals, and when they did, it was a small fraction of the total combinations tested^8,15,34^. By focusing on outcome-diet pairs with strong G×D effects and prioritizing interaction-specific SNPs, our study establishes an innovative approach to G×D risk prediction.

An important question for nutrigenomics is whether individualized dietary recommendations can be made based on G×D polygenic scores. We tested the hypothesis that unrealized G×D scores can stratify response to dietary exposure, focusing on alcohol consumption. Gout emerged as a particularly relevant example due to the well-established etiological role of alcohol in its development. Alcohol, particularly beer and spirits, is known to elevate serum uric acid by increasing purine metabolism and reducing renal urate excretion, thereby promoting gout onset in genetically susceptible individuals^35,36^. Our results indicate that individuals in the top quintile of unrealized G×D scores exhibited a strong susceptibility to alcohol with respect to gout risk (OR = 1.24 per additional alcoholic drink per day, *p* < 0.001) whereas individuals in the bottom quintile are relatively resistant (OR=1.04, *p*>0.05). Indeed, individuals with both high alcohol intake and high unrealized G×D scores had the highest risk of gout over 15 years of follow-up. As alcohol consumption is a modifiable lifestyle factor, this example provides a model for the potential of personalized dietary guidance.

Another major finding was the heritability estimates for the three categories of dietary exposures: SNIs, dPCs, and DIs. All exposures displayed a range of significant but modest heritability (range 5.0 to 8.8%). This is consistent with emerging data from other heritability methods used on UKB dietary data. Small cohort studies (< 2,000 individuals) have also found evidence for dietary intake heritability^37^. Furthermore, several GWASes have recently reported SNIs to be heritable, with *h*^2^ estimates ranging from 20-70%^38^. In the UKB, Cole et al., 2020^39^ found 84.1% of analyzed dietary habits (83/85 food-intake quantitative traits and 60/85 dPCs) were significantly heritable. Similarly, MonsterLM found all diet exposures to be modestly heritable and similar in *h*^2^ magnitude. For instance, dPC1 and alcohol intake had heritability of 0.085 and 0.088 versus 0.136 and 0.121 using alternative methods.

There are limitations to this study. Dietary data in the UKB is taken from FFQs and 24-hour recall questionnaire. Some drawbacks of this approach are memory biases, day-to-day variability, seasonal variations, and the likelihood of missing a rare diet event. Recent studies have commented on the impacts of self-reporting inaccuracy^40^ and imprecision in the UKB questionnaires^41^. Also, the 24-hour recall was taken as follow-up questions for UKB participants, while the FFQ was taken at baseline. Accordingly, SNI and DI diet variables used in this study have a sample size of approximately 141,000 individuals, while the dPCs had a sample size of up to 325,989 individuals, as they were derived from the FFQ. While MonsterLM offers numerous advantages compared to other methods for G×E estimation, the method has limitations. These include use of quantile normalization (which can bias estimates towards the null), potential information loss in variant QC, LD filtering, rare and low-frequency variant exclusion, and the standardization and liability adjustments in binary variables. For these reasons, null results may not reflect a lack of true G×D, but rather imprecision in the dietary variables or power loss due to steps in sample sizes filtering and/or methodology processing. As methodological approaches (such as phenotype integration^42^) continue to improve along with improvements in phenotyping within biobanks^43^, G×D power and detection will progress. Additionally, G×D scores themselves have inherent limitations. Their predictive performance depends on the strength of the underlying G×D effect, and weaker interactions may result in scores with limited clinical utility^44^. Moreover, G×D scores are context-dependent, meaning their applicability may vary across different populations, dietary environments, and genetic architectures. For instance, the variability in genetic backgrounds and environmental exposures can influence the effectiveness of G×D scores across diverse groups^45^. Further, while our study demonstrates their utility for disease risk stratification, the direct integration of G×D scores into clinical nutrition guidelines would require further validation in prospective dietary intervention studies.

In this report, we present a comprehensive study of genome-wide G×D in a biobank dataset across 3 major dietary classes and use these results to inform development of G×D scores. Our results expand the understanding of dietary classes and health outcomes contributing to G×D. We observed a specific pattern of genetic architecture for G×D, with evolutionarily recent dietary components (i.e. caffeine, alcohol) the most likely to show significant G×D. Moreover, most of the micro/macronutrients, DIs, and dPCs tested did not provide evidence of G×D. These findings guided the successful creation and application of G×D polygenic scores in complex traits. Altogether, this study helps lay the groundwork for future investigation in nutrigenomics, biobank-scale G×D genetic architecture quantification, and G×D polygenic score creation.

## Methods

### UK Biobank Genetic Data

The UKB is a prospective cohort study comprising over 500,000 participants residing in the United Kingdom with extensive phenotypic and genotypic data from men and women aged 40 to 69 collected between 2006 and 2010^46,47^ (https://www.ukbiobank.ac.uk/). Genotype information was processed using PLINK version 1.9 (https://www.cog-genomics.org/plink2/). A subset of 325,989 unrelated British individuals (comprising 54% female and 46% male) with available genotypic and phenotypic data were selected for the analysis. This unrelated cohort was chosen to mitigate potential inaccuracies in genomic predictions^48^. The sample size of 325,989 was further filtered to a subset of 141,144 individuals or less to ensure no missing data for 24-hour dietary recall data and outcomes for each outcome-diet combination. Exclusion criteria for individuals encompassed: (1) non-white British ancestry, (2) elevated ancestry-specific heterozygosity, (3) substantial genotype missingness (>0.05), (4) incongruent genetic ancestry, (5) sex chromosome aneuploidy, and (6) inconsistencies between gender sex and genetic sex, along with as well as consent withdrawal during analysis (**Extended Data Figure 1; Figure 1**). Variants from release version 3 of the UK Biobank data were used, including those found in the Haplotype Reference Consortium and 1000 Genomes panels. All variants had imputation quality exceeding 0.7 and no deviation from Hardy-Weinberg equilibrium (*P*>1×10^−10^)^46^. This study exclusively focused on common variants, leading to the filtering of genotypes by excluding highly correlated SNPs with an LD r^2^ > 0.9 and SNPs with a Minor Allele Frequency (MAF) < 0.05. SNP exclusion criteria involved: (1) SNPs with low imputation quality (INFO score < 0.30), (2) a call rate of less than 0.95, and (3) ambiguous or duplicated SNPs. Following the application of all quality control (QC) filters, 1,030,579 SNPs and 325,989 individuals were retained. Genetic variants were partitioned to minimize the number of blocks on each chromosome, with each block containing a maximum of 25,000 SNPs. All genotypes were standardized to a mean of zero and a standard deviation of one.

### UK Biobank Outcome and Diet Data

The MonsterLM G×D screen was applied to 31 outcomes (**Figure 2**) in the unrelated British participant cohort from the UKB. Outcomes that were tested included clinically relevant blood biomarkers, major diseases, and key anthropometric traits: BMI, WHR, and height. MonsterLM was applied with 23 dietary exposures consisting of three classes: 1) dPCs, 2) DIs, and 3) SNIs. dPCs were derived from the FFQ that was taken by most participants at baseline in the UKB. Using FFQ-based dPCs to describe dietary patterns and investigate G×D complements single-food intake approaches and can elicit stronger associations with dietary quality^39,49^. Furthermore, the high completion rates of the FFQ enable the analysis of up to 325,989 unrelated British participants, thus increasing power. dPCs were generated by applying the “prcomp” function in base R to standardized FFQ data with 17 food and drink features (**Extended Data Table 1**). These features included fruit and vegetables, meat, poultry, dairy, and salt. Variance explained for each dPC (out of 17 dPCs in total) and relative loadings were consistent with previously published FFQ-PCs derived from the UKB^49^ (**Extended Data Table 1**). G×D analyses were conducted using dPC1 and dPC2 as dietary pattern exposures.

The second class is DIs, dietary indexes, which were calculated through SNIs defined by total weight in metric units from the UKB’s online 24hr recall^50^ instances. The mean values of each SNI across all 24hour recall follow-up instances were used to calculate six major DIs using Zhan *et al.,* 2023’s validated *Dietaryindex* R package^51^. The DIs calculated included: DASH^52^, Healthy Eating Index-2020^53^, Alternative Healthy Eating Index (AHEI)^54^, MED Index in serving sizes from the PREDIMED trial (MEDI)^55^, Dietary Inflammatory Index (DII)^56^, American Cancer Society 2020 dietary index (ACS 2020)^57^.

The third class is SNIs which are represented by their total intake weight in metric units. The protocol to derive the weight of each nutrient group is described in Perez-Cornago, A. *et al.,* 2021’s nutrition calculation protocol from the Oxford WebQ questionnaire^50^. The Oxford WebQ is a user-friendly, automated, web-based dietary assessment tool that collects information about individuals’ food and drink consumption over the previous 24 hours, with nutrient estimation based on the UK Nutrient Databank (UKNDB). Mean values across all instances of the 24-hour recall were used for each SNI in six nutrient categories. Nutrient categories used included the following: carbohydrates (carbohydrates, fructose, and sucrose); proteins (protein and animal protein); fats (cholesterol, fat, n-6 fatty acids, and saturated fatty acids); minerals (sodium, potassium, and calcium); vitamins (vitamin A retinol equivalents); and discretionary items (alcohol and caffeinated coffee).

### Genome-wide Gene-Diet Interaction Analyses with MonsterLM

Genome-wide G×D estimation was conducted using MonsterLM (v0.1.1)^21^. Briefly, MonsterLM is a multiple linear regression-based method for estimation of trait variance explained by genotype and genotype-environment interactions. By leveraging multiple linear regression, MonsterLM provides estimates of adjusted R^2^ to assess heritability (*h*^2^) and variance of G×E interactions. MonsterLM efficiently calculates genetic and G×E effects without making prior assumptions in the genetic model (other than additive genetic effects) or variable parametrization. MonsterLM estimates the variance explained by G×D for *j* outcomes and *i* diet exposures by fitting multiple regression estimates to satisfy the linear model:

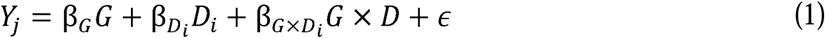

where *G* is the standardized genotype matrix, *D* is the quantile normalized diet exposure, *Gx D* is the product of the genotype matrix and diet exposure matrix, with the same dimensionality as *G*. MonsterLM estimates the total G×D estimate across 1,030,579 SNPs. Similarly, confidence intervals and *p*-values for each G×D estimate is calculated from the variance of the adjusted R^2^ ^21^. In this report, all instances of G×D estimation refer to the estimated adjusted R^2^ calculated through MonsterLM.

### G×D Score Analyses

To create G×D polygenic scores, we defined the following model:

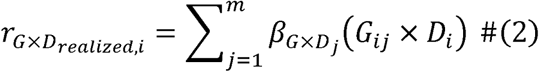

where *r_G×D_realized__* represents the G×D “realized G×D score”, *i* represents the individual number, *j* is the variant number, *β_G×D_j__* is the interaction (between outcome and diet exposure of interest) weight for each variant, *G_ij_* is the allele number for the *j^th^* variant of the *i^th^* individual, and *D_i_* is the dietary factor. For each outcome and diet exposure combination, participants were split into a discovery set (80%) and validation set (20%) where the discovery set was used to train outcome-diet specific interaction weights (*β_G×D_j__*), while the validation set, comprising non-overlapping participants, ensured independent evaluation. Those weights were then used to calculate the G×D score in the validation cohort. Five levels of *p*-value thresholding were used to select SNPs with a univariate association in the discovery set at levels: 0.25. 0.1, 0.05, 0.01, and 0.001. A clumping and threshold procedure was used with an LD *r*^2^ of 0.1 and clumping window size of 500 kb^27,58^. Realized G×D scores were regressed onto outcomes in the validation cohort.

Another related G×D score is the “unrealized G×D score.” The unrealized G×D score simply does not multiply with the corresponding diet exposure and is defined as:

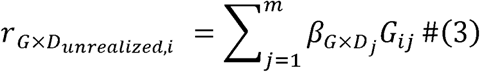

where *r_G×D_unrealized,i__* represents the unrealized G×D score without the diet environmental term *D_i_* (as seen in equation 5); *i* represents the individual number, *j* is the variant number, *β_G×D_j__* is the interaction (between outcome and diet exposure of interest) weight for each variant, and *G_ij_* is the allele number for the *j^th^* variant of the *i^th^* individual. All pertinent terms and definitions related to the MonsterLM G×D framework and G×D polygenic scores are provided in **Extended Data Table 10**.

Time-to-event analysis was conducted using a Cox regression model. Participants were stratified into four groups based on alcohol intake (Low vs. High) and unrealized G×D scores (above vs below median). Alcohol intake was categorized as Low for participants consuming alcohol ≤3 times per month and High for those consuming alcohol ≥3 times per week, excluding moderate drinkers. The resulting dataset was used to examine the association between alcohol intake and unrealized G×D scores with incident gout risk outcome over a 15-year follow-up period. Survival analysis was performed using the ‘coxph’ function in R, with an interaction term between alcohol intake and unrealized G×D score to evaluate the combined effect of alcohol consumption and G×D polygenic score on gout incidence. The interaction model assessed whether the effect of alcohol consumption on gout risk differs depending on the level of the G×D score. The statistical significance of the interaction term was assessed using likelihood ratio tests, comparing the full model (with the interaction) to a reduced model (without the interaction). A *p*-value of less than 0.05 was considered statistically significant.

### Phenome-wide Association Study with G×D Scores

A PheWAS was performed to investigate associations between realized G×D scores, derived from the alcohol diet variable, and ICD-10 phecodes in a validation subset of unrelated British individuals from the UK Biobank cohort. Phecodes with a minimum number of cases, 100, were included. The primary independent variable in the analysis was the realized G×D score, which captures the interaction effect of genetic variants and diet. Logistic regression was used to assess the association of the realized G×D score with each binary phecode phenotype. Adjustments for age, sex, and genetic ancestry, as measured by principal components, were applied as covariates to account for potential confounding and population structure. For each phecode, the odds ratio (OR) and 95% confidence intervals were calculated to quantify the association between the realized G×D score and the likelihood of having the phenotype. Bonferroni correction was applied to adjust *p*-values, controlling for type I error.

## Data Availability

This research was conducted using individual-level data from the UK Biobank (https://www.ukbiobank.ac.uk/
) under application #15255. The UK Biobank data are not publicly available but can be accessed by bona fide researchers upon application. All other data supporting the findings of this study are available within the article and its Supplementary Information files.

https://github.com/GMELab/MonsterLM.

## Acknowledgements

The authors are thankful for all the UK Biobank participants. We would also like to thank Dr. Andrew McArthur and CSU at McMaster University for providing computing support. We thank James Feiner for UKB and software support.

M.D. is supported by a Canadian Institute of Health Research Doctoral Award.

## Author Contributions

**MD**: conceptualization, software, formal analysis, investigation, visualization, writing; **AM, RL, JW, AL, PF**: formal analysis, writing (review and edit); **GP**: conceptualization, supervision, funding acquisition, methodology, project administration, writing (review and edit).

## Ethic Approval, Data Availability, and Code

This research was conducted using individual-level genetic and phenotypic data from the UK Biobank under application #15255. The UK Biobank study received ethical approval from the National Health Service National Research Ethics Service North West Multi-centre Research Ethics Committee (REC reference: 11/NW/0382), and all participants provided written informed consent.

All analyses were performed in accordance with relevant guidelines and regulations, and in compliance with UK Biobank data access protocols.

UK Biobank data are available to registered researchers upon application (https://www.ukbiobank.ac.uk/). The individual-level data used in this study are not publicly available due to UK Biobank access restrictions but can be accessed through the UK Biobank resource under the appropriate application. All other data supporting the findings of this study are available within the article and its Supplementary Information files.

Code used for the analyses is available at: https://github.com/GMELab/MonsterLM.

## Extended Data Figures

**Extended Data Figure 1.**
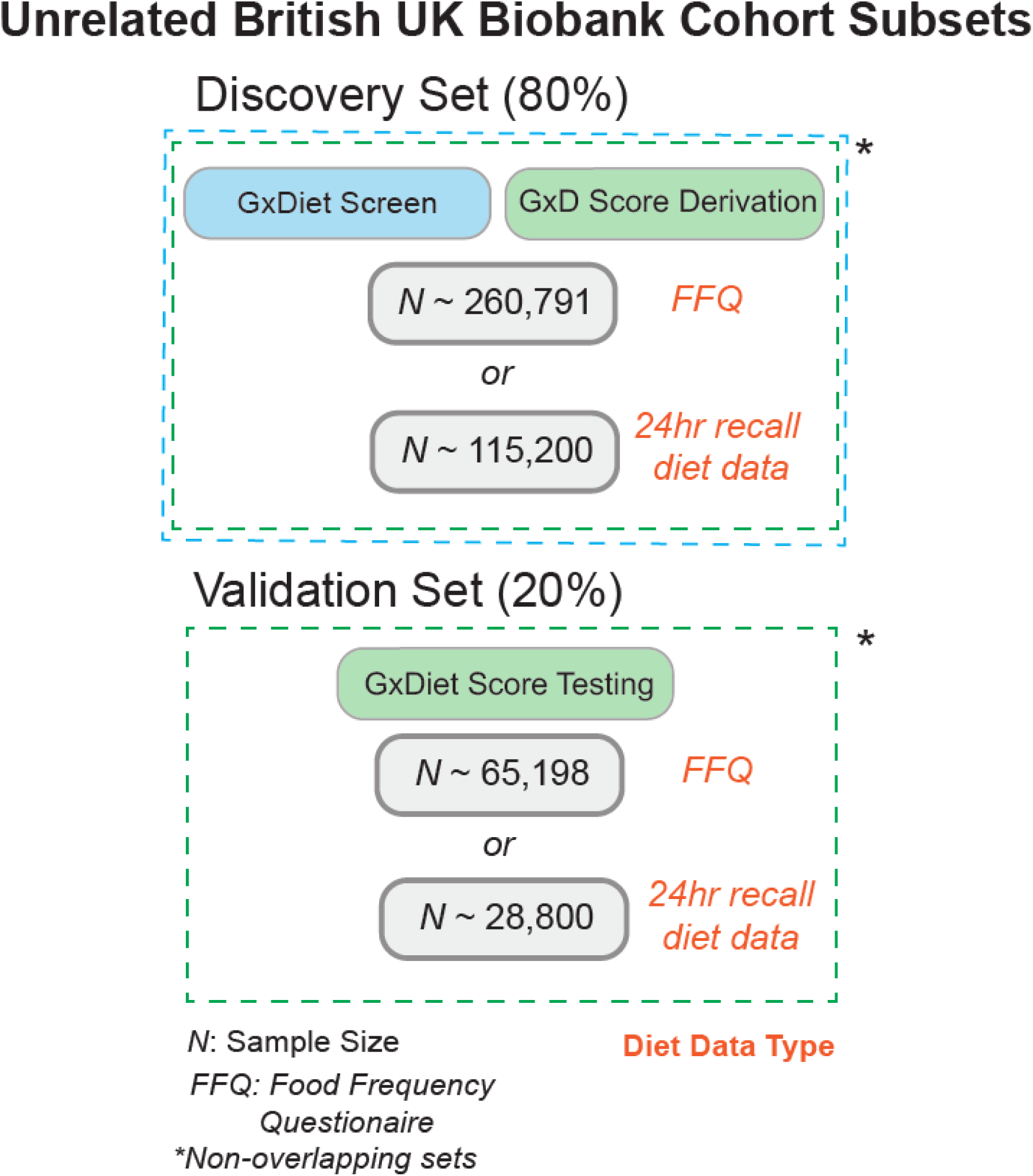
| Overview of UK Biobank cohort subsets for G×D analysis. The discovery set (80%) consists of two groups: 1) approximately 260,791 participants with data from the Food Frequency Questionnaire (FFQ), or 2) approximately 115,200 participants with data from the 24-hour recall diet. These cohorts were used for the G×D screen and G×D score derivation. The validation set (20%) includes either approximately 65,198 participants with FFQ data or approximately 28,800 participants with 24-hour recall diet data. These subsets were used for G×D score testing. All cohorts are non-overlapping, and the participant count may slightly vary based on the MonsterLM protocol used for selection and analysis.

**Extended Data Figure 2.**
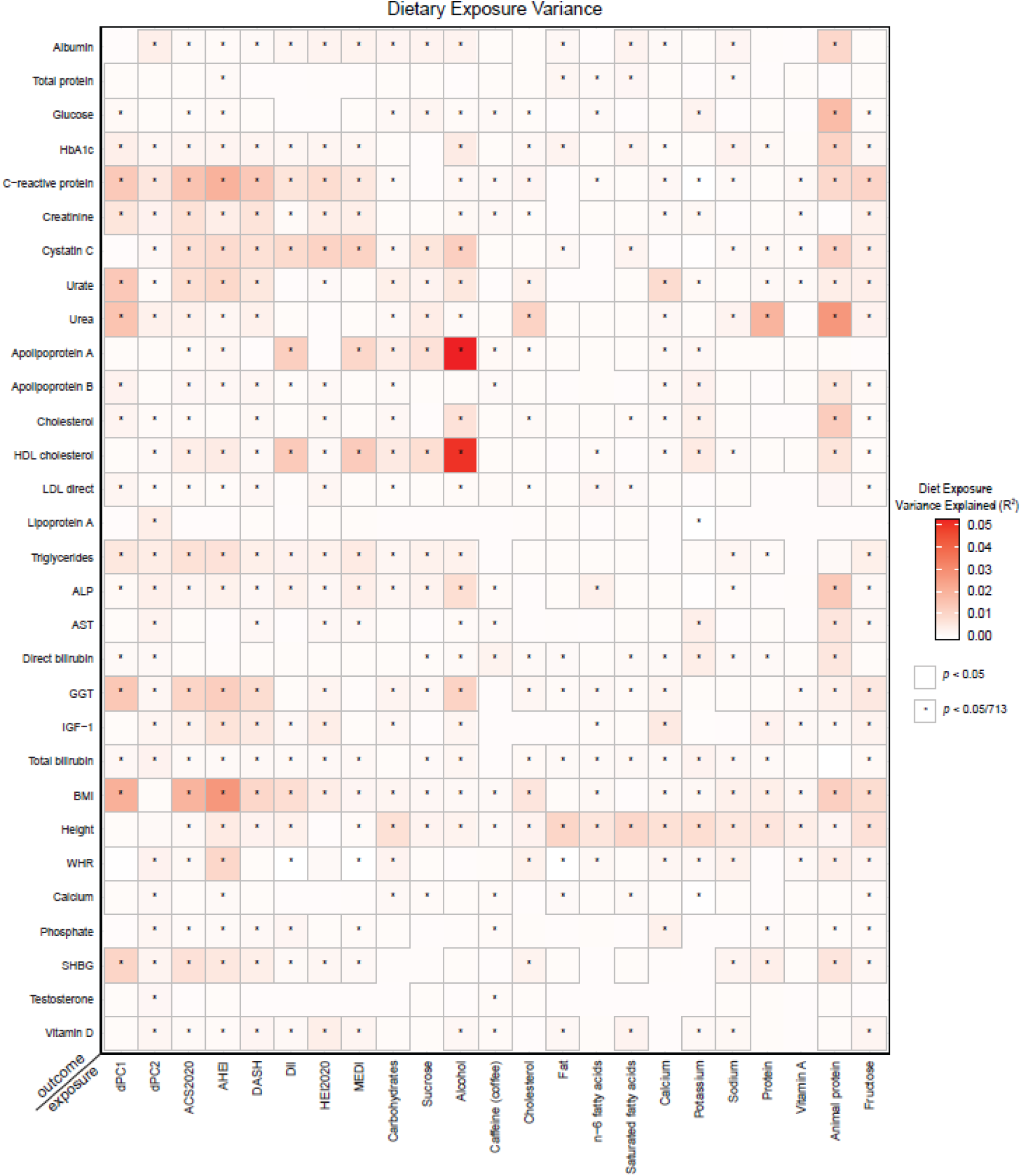
| Estimates of dietary exposure variance across 23 dietary exposures and 31 health outcomes. Estimates of the outcome variance explained by the exposure only. Outcomes are on the y-axis and dietary exposures are on the x-axis. Each cell contains the total exposure variance (adjusted R^2^) estimate. Cells outlined in grey are nominally significant (*p*<0.05) and cells that are marked with an asterisk are Bonferroni significant (*p*<0.05/713).

**Extended Data Figure 3.**
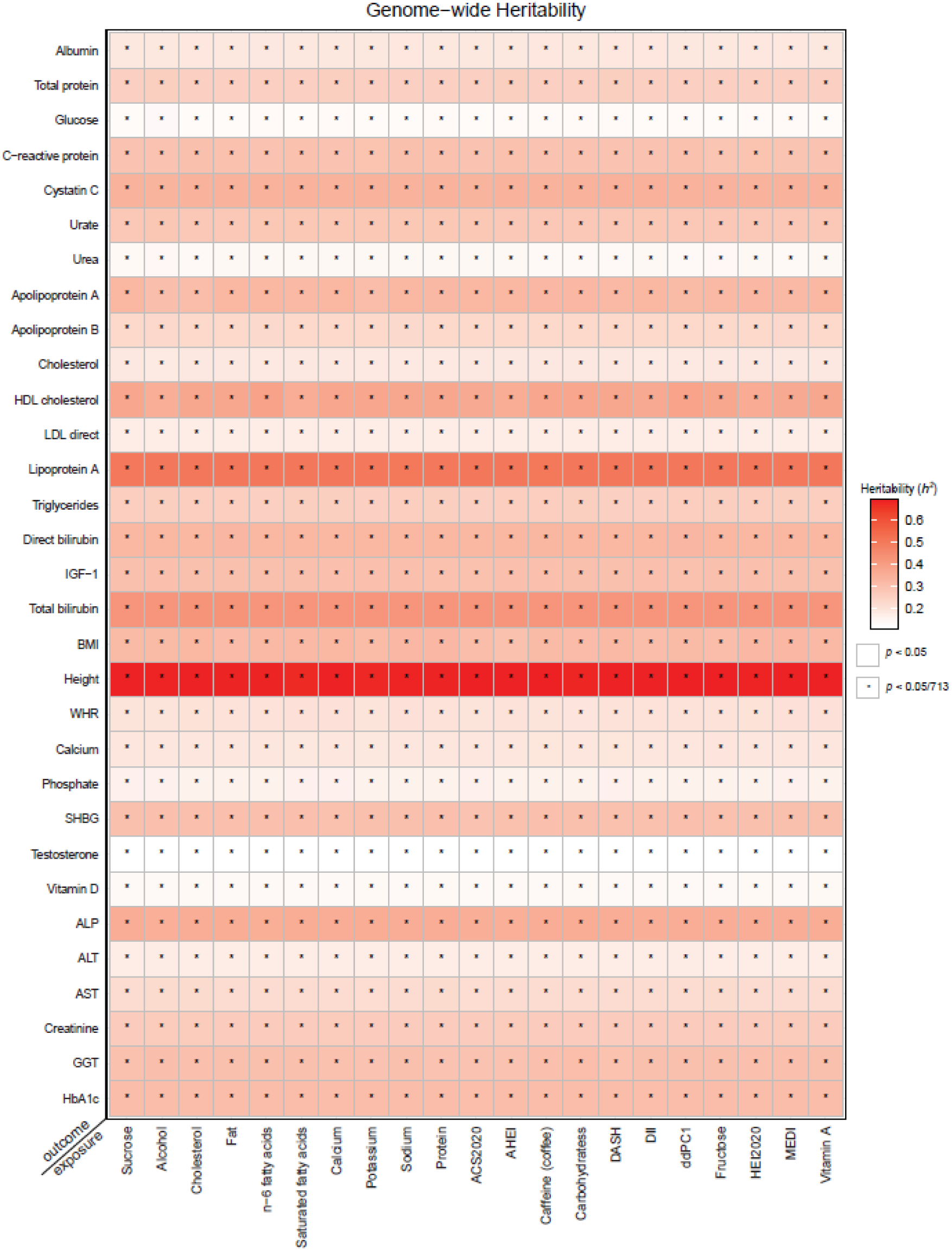
| Estimates of heritability (*h*^2^) across 23 dietary exposures and 31 traits and biomarker outcomes. Estimates computed using the MonsterLM heritability methodology. Outcomes are on the y-axis and dietary exposures are on the x-axis. Each cell contains the total *h*^2^ estimate. Cells outlined in black are nominally significant (*p*<0.05) and cells that are marked with an asterisk are Bonferroni significant (*p*<0.05/713).

**Extended Data Figure 4.**
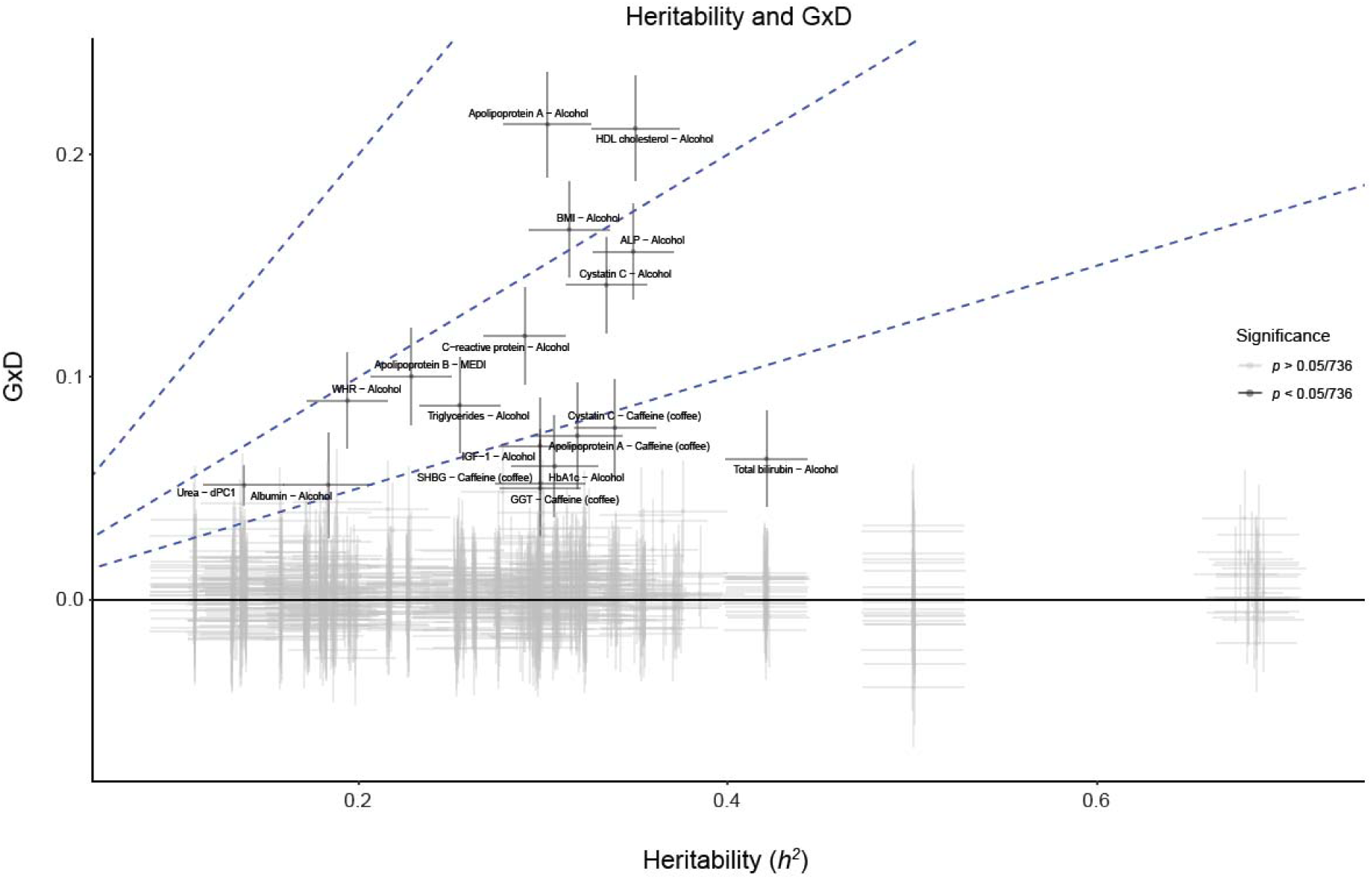
| Heritability compared to G×D for every outcome and exposure combination. Blue sloped lines represent y=x, y=0.5x, and y=0.25x to illustrate the proportion of heritability to G×D for each combination tested. Horizontal and vertical error bars are 95% confidence intervals for heritability and G×D estimates, respectively. Bonferroni significant estimates are defined as *p*<0.05/713 and are coloured black with labels.

**Extended Data Figure 5.**
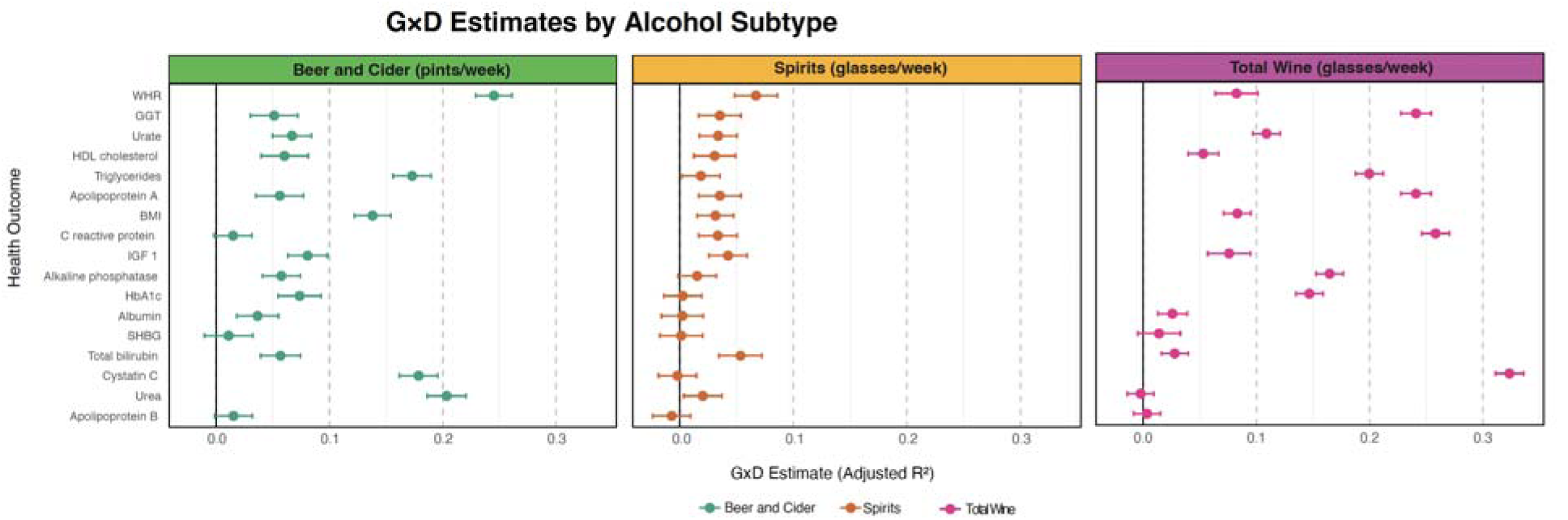
| G×D estimates by alcohol subtype. Forest plot of G×D estimates from the discovery cohort across four alcohol diet categories: beer and cider (pints/week), spirits (glasses/week), and total wine (glasses/week). Each point represents the adjusted R² (G×D estimate) from MonsterLM for a specific health outcome (y-axis), with horizontal lines indicating the 95% confidence intervals.

**Extended Data Figure 6.**
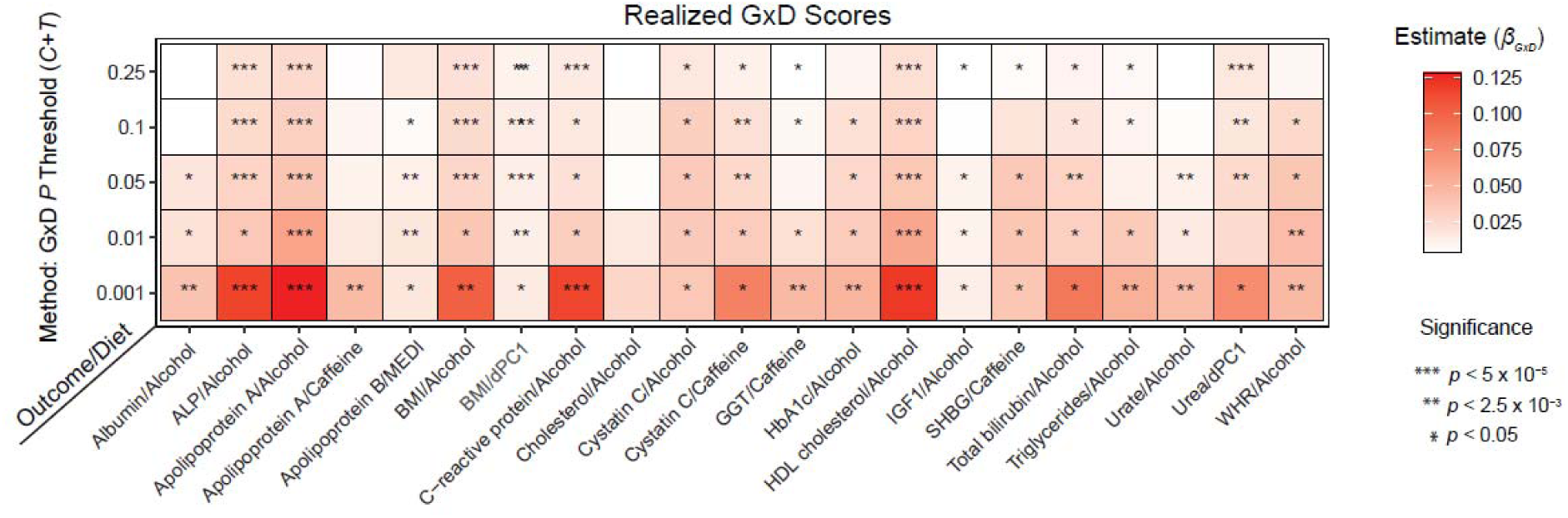
| Realized G×D Score Estimates Associated with a validation cohort. Estimates ( ) represent the regression coefficient from the validation cohort realized G×D score regressed onto the cohort’s outcome value. The colour gradient ranges from light red to dark, indicating the magnitude of the regression coefficient. Significance levels are indicated by asterisks representing two levels of significance: * as nominally significant (p<0.05) and ** as Bonferroni significant (p<0.05/20). The horizontal axis displays outcome-diet pairs, and the vertical axis displays the SNP selection type model based on the univariate G×D *P* value threshold from the discovery set. Validation cohort size was 20% that of the sample size used in the main screen (N∼28,000 per combination). G×D PRS was generated using the *p*-value thresholding approach.

**Extended Data Figure 7.**
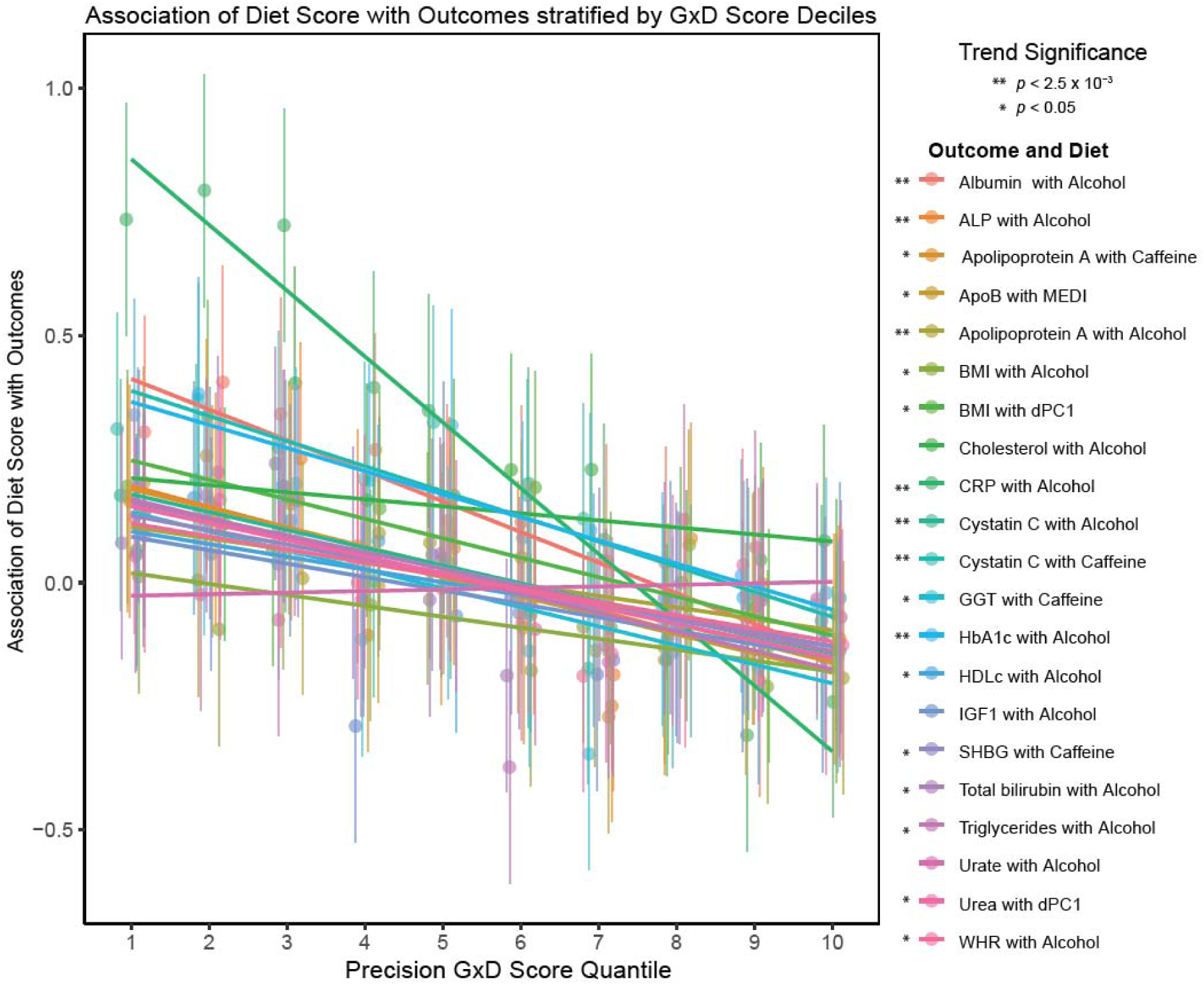
| Precision Diet Estimates. Precision Diet estimates across 20 identified G×D score combinations. The x-axis represents the deciles of the precision G×D scores, while the y-axis shows the predicted diet scores, calculated by weighting the diet variables using coefficients estimated from the discovery set. Trends and significance are depicted through simple linear regression of the quantiles on diet scores. The 20 significant G×D combinations were selected from the initial screen, along with a negative control (Cholesterol with Alcohol). Error bars at each quantile indicate 95% confidence intervals based on observed diet score data. Significance is denoted by asterisks: * for nominal significance (p < 0.05) and ** for Bonferroni-adjusted significance (p < 0.05/20). The validation cohort size was approximately 20% of the main sample used in the initial screen (N ∼ 28,000 per combination). G×D scores were derived using a *p*-value thresholding approach.

## Extended Data Tables

**Extended Data Table 1.** Correlation Matrix for all Dietary Index Scores (Pearson’s Correlation Coefficient)

| | DASH | HEI2020 | AHEI | MEDI | DII | ACS2020 | *PCC* ($\vert \bar{r}\vert $) |
| --- | --- | --- | --- | --- | --- | --- | --- |
| DASH | 1 | 0.532835 | 0.654367 | 0.363959 | -0.23975 | 0.616938 | 0.481569 |
| HEI2020 | 0.532835 | 1 | 0.44912 | 0.450454 | -0.24743 | 0.624192 | 0.460806 |
| AHEI | 0.654367 | 0.44912 | 1 | 0.365668 | -0.33303 | 0.736704 | 0.507777 |
| MEDI | 0.363959 | 0.450454 | 0.365668 | 1 | -0.30229 | 0.31545 | 0.359565 |
| DII | -0.23975 | -0.24743 | -0.33303 | -0.30229 | 1 | -0.20216 | 0.264931 |
| ACS2020 | 0.616938 | 0.624192 | 0.736704 | 0.31545 | -0.20216 | 1 | 0.499088 |

**Dietary Indexes Pearson's Correlation Matrix.** A Pearson correlation coefficient (PCC) score matrix for each vector of dietary index scores compared to another. The PCC column denotes the average PCC magnitude (excluding a direct comparison to the same dietary index) per dietary index. 141,144 individuals used to calculate scores.

**Extended Data Table 2.** Contribution of each Dietary Trait to the Principal Component Characteristics from Food Frequency Questionnaire.

| Dietary Pattern Principal Components derived from the Food Frequency Questionnaire |  |  |  |  |  |
| --- | --- | --- | --- | --- | --- |
| FFQ Feature<br>(PC Variance Explained) | dPCs Loadings (variance explained) |  |  |  |  |
|  | dPC1<br>(13.5%) | dPC2<br>(9.9%) | dPC3<br>(7.2%) | dPC4<br>(6.6%) | dPC5<br>(6.5%) |
| Cooked vegetable intake | -0.01547 | 0.295203 | -0.02951 | 0.226142 | -0.26987 |
| Fresh fruit intake | -0.12589 | 0.377397 | 0.019578 | 0.268383 | 0.013468 |
| Oily fish intake | 0.028967 | 0.529346 | -0.26844 | -0.23882 | -0.12218 |
| Non-oily fish intake | 0.105807 | 0.458744 | -0.3324 | -0.30099 | -0.13354 |
| Processed meat intake | 0.415316 | -0.17933 | -0.20467 | -0.06562 | 0.043483 |
| Poultry intake | 0.338158 | 0.198788 | 0.0984 | -0.06983 | 0.099493 |
| Beef intake | 0.466133 | 0.009606 | 0.139873 | 0.121797 | 0.081175 |
| Lamb/mutton intake | 0.455591 | 0.104237 | 0.125326 | 0.114031 | 0.049395 |
| Pork intake | 0.463373 | 0.048205 | 0.097654 | 0.125275 | 0.069704 |
| Never eat eggs, dairy, wheat, sugar | 0.065635 | -0.16742 | -0.23659 | -0.13321 | 0.194992 |
| Cheese intake | 0.029876 | -0.17983 | -0.43355 | -0.14905 | 0.077905 |
| Bread intake | 0.100553 | -0.20237 | -0.56949 | 0.093756 | -0.03773 |
| Cereal intake | -0.08591 | 0.186048 | -0.11267 | 0.365052 | 0.522189 |
| Salt added to food | 0.132334 | -0.22405 | 0.037819 | -0.11231 | -0.5048 |
| Tea intake | 0.041637 | -0.00665 | -0.30166 | 0.525733 | -0.26795 |
| Hot drink temperature | 0.000684 | 0.068711 | 0.025348 | -0.41027 | 0.364949 |
| Variation in diet | 0.059541 | 0.062601 | 0.213977 | -0.19271 | -0.30092 |

**Principal Component Descriptive Characteristics.** Genome-wide G×D estimates were performed across 325,989 unrelated British individuals from the UKB. The 17 FFQ features used to create the dPCs are displayed. For each of the first 5 dPCs, the relative loading weight and the proportion of total variance explained by that dPC as a fraction of the variance explained by all PCs is shown.

**Extended Data Table 3a.** Top 20 G×D Estimates from the Whole Cohort.

| Health Outcome | Diet Exposure | G×D Estimate (p) | Diet Class |
| --- | --- | --- | --- |
| Apolipoprotein A | Alcohol | 0.213 ($1.02 \times 10^{-63}$) | Single Nutrient Intake |
| HDL cholesterol | Alcohol | 0.212 ($2.29 \times 10^{-63}$) | Single Nutrient Intake |
| BMI | Alcohol | 0.166 ($2.45 \times 10^{-46}$) | Single Nutrient Intake |
| Alkaline phosphatase | Alcohol | 0.156 ($1.30 \times 10^{-41}$) | Single Nutrient Intake |
| Cystatin C | Alcohol | 0.141 ($1.08 \times 10^{-34}$) | Single Nutrient Intake |
| Urea | dPC1 | 0.051 ($4.28 \times 10^{-27}$) | Dietary Pattern PC |
| C-reactive protein | Alcohol | 0.118 ($5.94 \times 10^{-25}$) | Single Nutrient Intake |
| Apolipoprotein B | MEDI | 0.100 ($1.80 \times 10^{-18}$) | Diet Index |
| WHR | Alcohol | 0.089 ($2.90 \times 10^{-15}$) | Single Nutrient Intake |
| Triglycerides | Alcohol | 0.087 ($1.07 \times 10^{-14}$) | Single Nutrient Intake |
| Cystatin C | Caffeine (coffee) | 0.077 ($7.07 \times 10^{-12}$) | Single Nutrient Intake |
| IGF-1 | Alcohol | 0.069 ($8.61 \times 10^{-10}$) | Single Nutrient Intake |
| Apolipoprotein A | Caffeine (coffee) | 0.074 ($2.88 \times 10^{-9}$) | Single Nutrient Intake |
| Total bilirubin | Alcohol | 0.063 ($1.56 \times 10^{-8}$) | Single Nutrient Intake |
| HbA1c | Alcohol | 0.060 ($3.02 \times 10^{-7}$) | Single Nutrient Intake |
| BMI | dPC1 | 0.021 ($3.34 \times 10^{-6}$) | Dietary Pattern PC |
| GGT | Caffeine (coffee) | 0.050 ($6.24 \times 10^{-6}$) | Single Nutrient Intake |
| Urate | Alcohol | 0.048 ($1.21 \times 10^{-5}$) | Single Nutrient Intake |
| SHBG | Caffeine (coffee) | 0.052 ($2.05 \times 10^{-5}$) | Single Nutrient Intake |
| Albumin | Alcohol | 0.052 ($2.22 \times 10^{-5}$) | Single Nutrient Intake |

**Top 20 G×D estimates with MonsterLM with the whole cohort.** Genome-wide G×D estimates were obtained across 141,144 to 325,989 unrelated British individuals from the UKB (whole available cohort). Estimates for the highest 20 G×D sorted by $p$ -values are displayed with their estimated adjusted $R^2$ . The larger dietary class for each outcome-diet combination is described in the right column. MEDI: MED Index in serving sizes from the PREDIMED trial. $p$ -values are from the two-tailed significance testing described in MonsterLM.

**Extended Data Table 3b.**
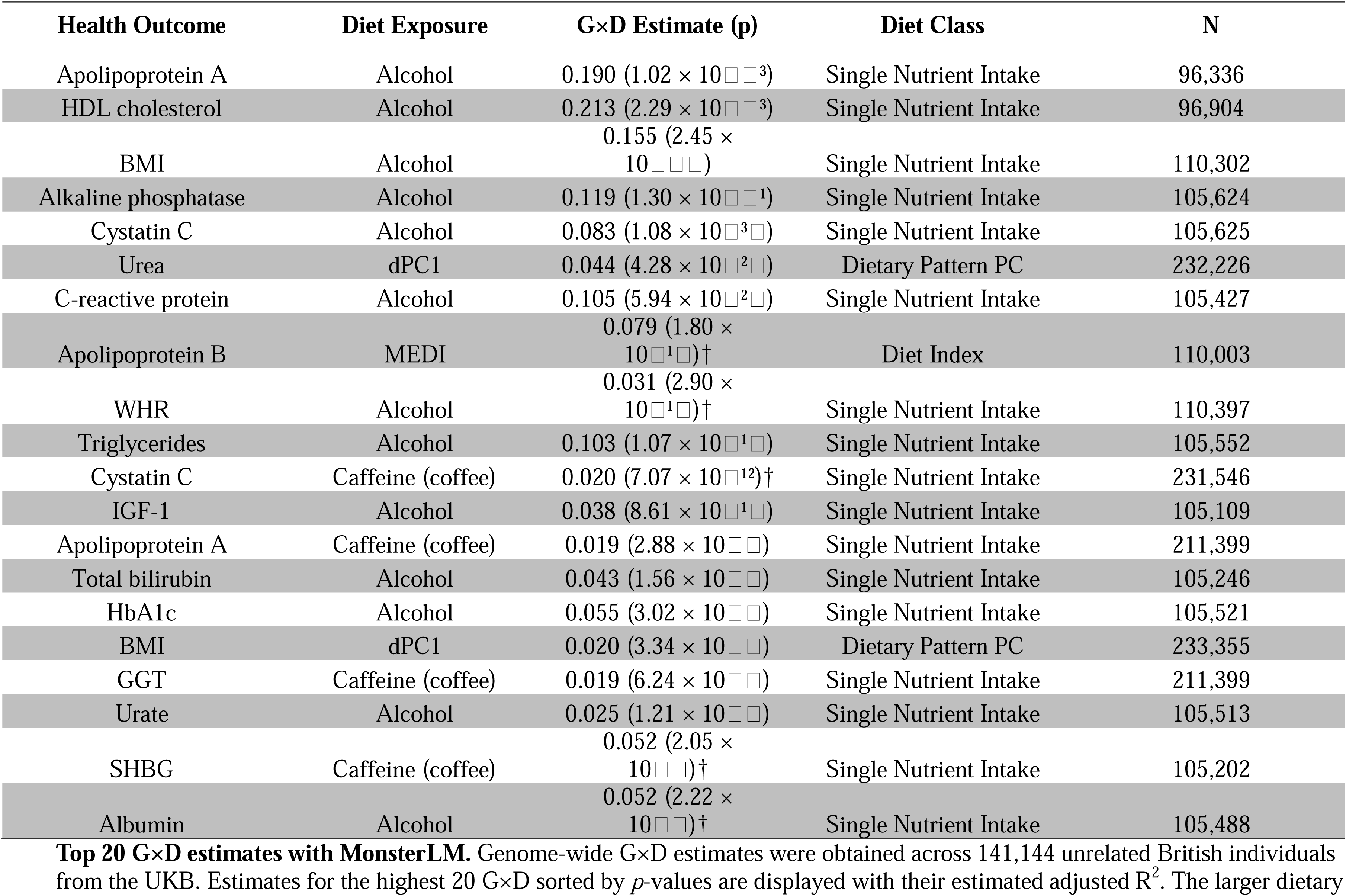
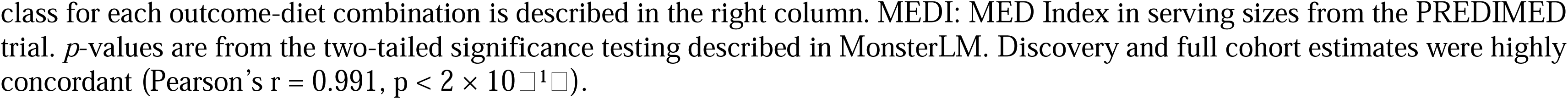
Top 20 G×D Estimates from the Discovery Cohort.

**Extended Data Table 4.** Apolipoprotein B with MEDI G×D with Lipid-lowering Medication Adjustments.

| Medication Label | G×D Estimate (95% CI) |  |
| --- | --- | --- |
|  | Medication Effect Adjustment | Medication Participant Removal |
| Omega 3 Fish Oil | 0.100 (0.079, 0.122) | 0.099 (0.077, 0.120) |
| Bezafibrate | 0.099 (0.078, 0.121) | 0.101 (0.079, 0.123) |
| Fenofibrate | 0.100 (0.079, 0.122) | 0.100 (0.078, 0.122) |
| Simvastatin | 0.099 (0.078, 0.121) | 0.047 (0.023, 0.071) |
| Fluvastatin | 0.098 (0.077, 0.120) | 0.096 (0.074, 0.117) |
| Pravastatin | 0.100 (0.079, 0.122) | 0.097 (0.076, 0.119) |
| Cod Liver Oil Capsules | 0.100 (0.079, 0.122) | 0.090 (0.068, 0.112) |
| Atorvastatin | 0.100 (0.079, 0.122) | 0.097 (0.075, 0.118) |
| Rosuvastatin | 0.101 (0.079, 0.123) | 0.100 (0.078, 0.122) |
| Ezetimibe | 0.101 (0.079, 0.123) | 0.099 (0.077, 0.120) |

**G×D estimate for Apolipoprotein B with MEDI across all lipid-lowering medication adjustments.** Genome-wide G×D estimates were obtained in up to 141,144 unrelated British individuals from the UKB. Estimates for the highest 20 G×D are reported with its estimated adjusted $R^2$ and 95% confidence intervals. Medication Effect Adjustment method computes the G×D after the raw Apolipoprotein B levels is adjusted for their medication in users using a meta-analyzed effect estimate unique to each drug. Medication participant removal method computes the G×D after users of the medication is removed from the analysis. MEDI: MED Index in serving sizes from the PREDIMED trial. $p$ -values are from the two-tailed significance testing described in MonsterLM. Sample size ranges from 126,105 to 140,383.

**Extended Data Table 5.**
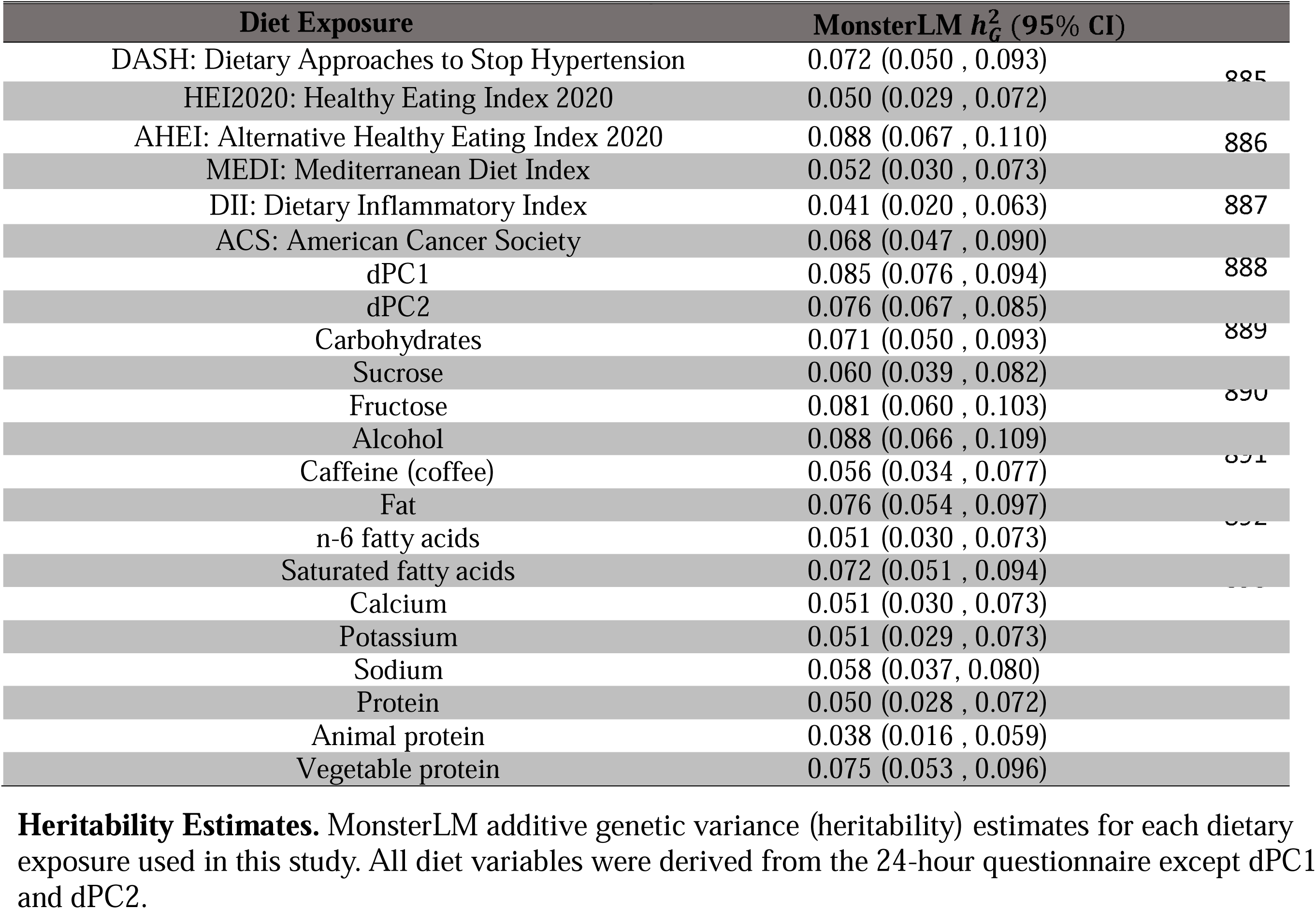
MonsterLM heritability for each Dietary Index Score, dPC, and SNI.

**Extended Data Table 6.** Association of unrealized G×D scores with their corresponding diet and outcomes.

| Outcome | Diet | Diet and G×D Score ($R^2$, $p$) | Outcome and G×D Score ($R^2$, $p$) |
| --- | --- | --- | --- |
| Apolipoprotein A | Alcohol | <0.001 , 0.081 | <0.001 , 0.478 |
| HDL cholesterol | Alcohol | <0.001 , 0.801 | <0.001 , 0.169 |
| BMI | Alcohol | <0.001 , 0.543 | <0.001 , 0.259 |
| Alkaline phosphatase | Alcohol | <0.001 , 0.393 | <0.001 , 0.220 |
| Cystatin C | Alcohol | <0.001 , 0.366 | <0.001 , 0.475 |
| Urea | dPC1 | <0.001 , 0.517 | <0.001 , 0.789 |
| C-reactive protein | Alcohol | <0.001 , 0.513 | <0.001 , 0.348 |
| Apolipoprotein B | MEDI | <0.001 , 0.247 | <0.001 , 0.328 |
| WHR | Alcohol | <0.001 , 0.325 | <0.001 , 0.902 |
| Triglycerides | Alcohol | <0.001 , 0.407 | <0.001 , 0.604 |
| Cystatin C | Caffeine (coffee) | <0.001 , 0.302 | <0.001 , 0.455 |
| IGF-1 | Alcohol | <0.001 , 0.224 | <0.001 , 0.238 |
| Apolipoprotein A | Caffeine (coffee) | <0.001 , 0.453 | <0.001 , 0.362 |
| Total bilirubin | Alcohol | <0.001 , 0.348 | <0.001 , 0.157 |
| HbA1c | Alcohol | <0.001 , 0.441 | <0.001 , 0.326 |
| BMI | dPC1 | <0.001 , 0.325 | <0.001 , 0.494 |
| GGT | Caffeine (coffee) | <0.001 , 0.585 | <0.001 , 0.064 |
| Urate | Alcohol | <0.001 , 0.395 | <0.001 , 0.505 |
| SHBG | Caffeine (coffee) | <0.001 , 0.569 | <0.001 , 0.474 |
| Albumin | Alcohol | <0.001 , 0.302 | <0.001 , 0.551 |

**Association of G×D scores with their corresponding diet and outcomes.** Diet and outcome variables were regressed onto unrealized G×D scores at a discovery $p$ -value threshold of 0.001. Regression adjusted $R^2$ and $p$ values are reported per outcome and diet exposure combination. Validation cohort size was 20% that of the sample size used in the main screen (N~28,000 per combination).

**Extended Data Table 7.** PheWAS of G×D scores (equation 6) with ICD-10 phecodes.

| Phecode | Phenotype | Category | Case/Control Count | Odds Ratio (OR) | P-value | Adjusted P-Value (Bonferroni) | Outcome | Diet | 95% Confidence Intervals |
| --- | --- | --- | --- | --- | --- | --- | --- | --- | --- |
| 274 | Gout and other crystal arthropathies | endocrine metabolic | 227/21532 | 0.984 | 0.276 | $1.07 \times 10^{-4}$ | BMI | Alcohol | 0.834, 1.161 |
| 550 | Abdominal hernia | digestive | 2647/19107 | 1.058 | 0.614 | $1.07 \times 10^{-4}$ | Alkaline phosphatase | Alcohol | 0.956, 1.17 |
| 535 | Gastritis and duodenitis | digestive | 1602/19942 | 0.969 | 0.546 | $1.07 \times 10^{-4}$ | Alkaline phosphatase | Alcohol | 0.855, 1.097 |
| 550.2 | Diaphragmatic hernia | digestive | 1503/19107 | 1.04 | 0.756 | $1.07 \times 10^{-4}$ | Alkaline phosphatase | Alcohol | 0.915, 1.183 |
| 599 | Other symptoms or disorders of the urinary system | genitourinary | 1236/18550 | 0.87 | 0.374 | $1.13 \times 10^{-4}$ | Apolipoprotein A | Alcohol | 0.361, 2.096 |
| 411.4 | Coronary atherosclerosis | circulatory system | 1048/18161 | 0.643 | 0.594 | $1.13 \times 10^{-4}$ | Apolipoprotein A | Alcohol | 0.243, 1.702 |
| 411.8 | Other chronic ischemic heart disease unspecified | circulatory system | 1312/18161 | 0.788 | 0.848 | $1.13 \times 10^{-4}$ | Apolipoprotein A | Alcohol | 0.327, 1.897 |
| 317.1 | Alcoholism | mental disorders | 489/20163 | 0.870 | 0.762 | $1.08 \times 10^{-4}$ | C reactive protein | Alcohol | 0.353, 2.14 |
| 411 | Ischemic Heart Disease | circulatory system | 1746/19932 | 1.048 | 0.854 | $1.08 \times 10^{-4}$ | C reactive protein | Alcohol | 0.632, 1.74 |
| 250.2 | Type 2 diabetes | endocrine metabolic | 967/18886 | 0.925 | 0.783 | $1.13 \times 10^{-4}$ | HDL cholesterol | Alcohol | 0.534, 1.603 |
| 250 | Diabetes mellitus | endocrine metabolic | 1031/18886 | 0.884 | 0.649 | $1.13 \times 10^{-4}$ | HDL cholesterol | Alcohol | 0.519, 1.51 |

**G×D PheWAS Estimates.** The association between various ICD-10 phenotypes and the G×D score (with denoted outcome and exposures) as the independent variable. The *p*-values were adjusted for multiple comparisons depending on the ICD-10 count used in each pheWAS. The G×D scores do not contain an interaction term, but rather the genetic data as per equation 6. Unrealized G×D scores of $p < 0.001$ thresholding was used.

**Extended Data Table 8.**
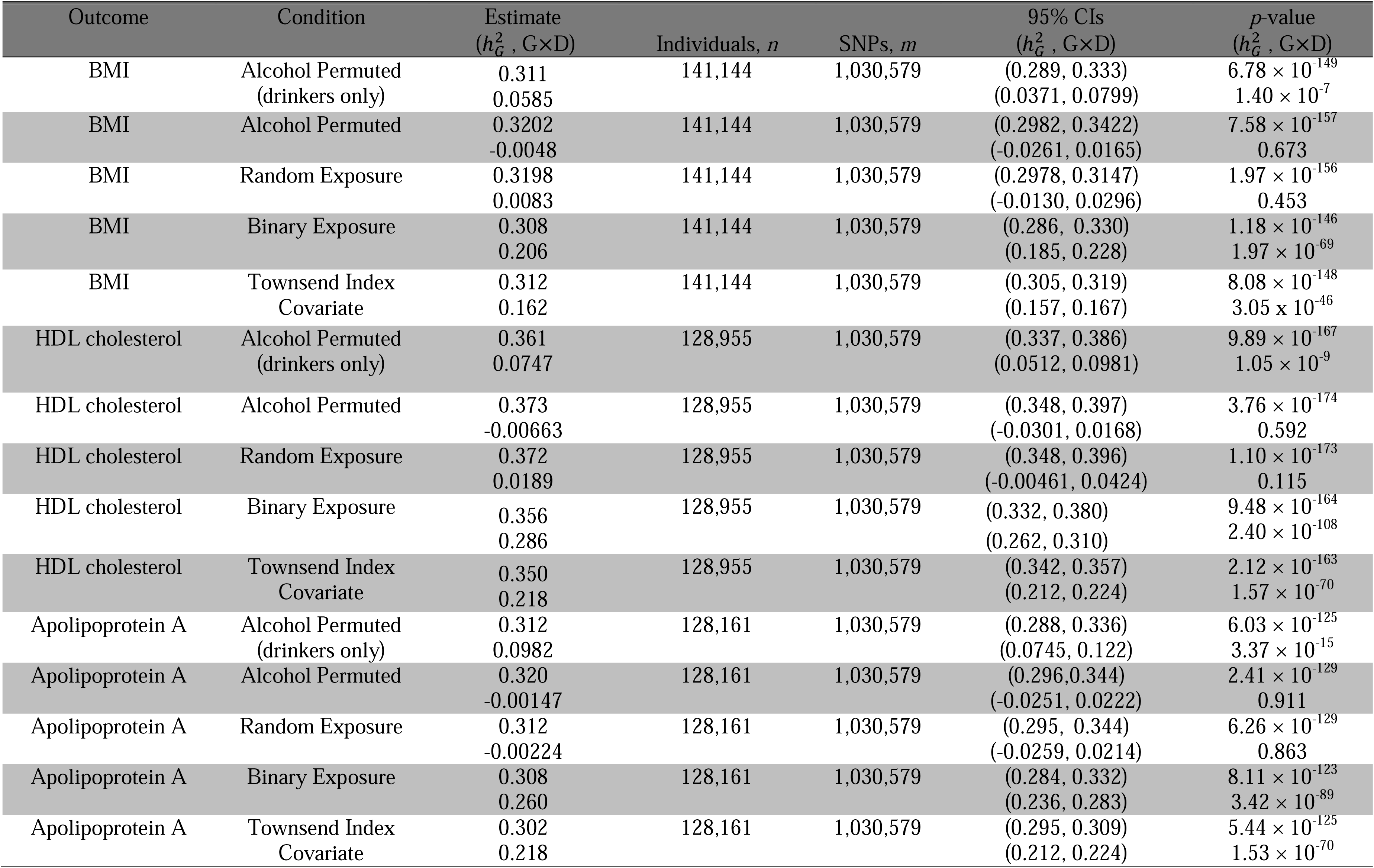
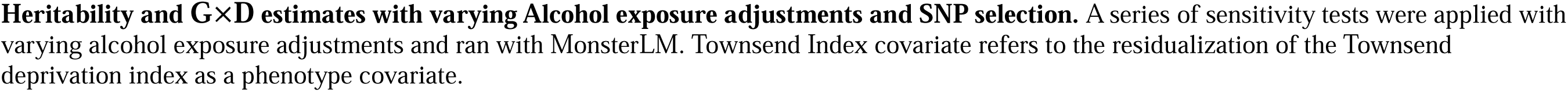
Sensitivity Analyses for BMI, HDL-cholesterol, and Apolipoprotein A with Alcohol.

| Outcome | Condition | Estimate<br>($h_G^2$, G×D) | Individuals, $n$ | SNPs, $m$ | 95% CIs<br>($h_G^2$, G×D) | $p$-value<br>($h_G^2$, G×D) |
| --- | --- | --- | --- | --- | --- | --- |
| BMI | Alcohol Permuted<br>(drinkers only) | 0.311 | 141,144 | 1,030,579 | (0.289, 0.333) | $6.78 \times 10^{-149}$ |
| | | 0.0585 | | | (0.0371, 0.0799) | $1.40 \times 10^{-7}$ |
| BMI | Alcohol Permuted | 0.3202 | 141,144 | 1,030,579 | (0.2982, 0.3422) | $7.58 \times 10^{-157}$ |
|  |  | -0.0048 |  |  | (-0.0261, 0.0165) | 0.673 |
| BMI | Random Exposure | 0.3198 | 141,144 | 1,030,579 | (0.2978, 0.3147) | $1.97 \times 10^{-156}$ |
|  |  | 0.0083 |  |  | (-0.0130, 0.0296) | 0.453 |
| BMI | Binary Exposure | 0.308 | 141,144 | 1,030,579 | (0.286, 0.330) | $1.18 \times 10^{-146}$ |
| | | 0.206 | | | (0.185, 0.228) | $1.97 \times 10^{-69}$ |
| BMI | Townsend Index<br>Covariate | 0.312 | 141,144 | 1,030,579 | (0.305, 0.319) | $8.08 \times 10^{-148}$ |
| | | 0.162 | | | (0.157, 0.167) | $3.05 \times 10^{-46}$ |
| HDL cholesterol | Alcohol Permuted<br>(drinkers only) | 0.361 | 128,955 | 1,030,579 | (0.337, 0.386) | $9.89 \times 10^{-167}$ |
| | | 0.0747 | | | (0.0512, 0.0981) | $1.05 \times 10^{-9}$ |
| HDL cholesterol | Alcohol Permuted | 0.373 | 128,955 | 1,030,579 | (0.348, 0.397) | $3.76 \times 10^{-174}$ |
|  |  | -0.00663 |  |  | (-0.0301, 0.0168) | 0.592 |
| HDL cholesterol | Random Exposure | 0.372 | 128,955 | 1,030,579 | (0.348, 0.396) | $1.10 \times 10^{-173}$ |
|  |  | 0.0189 |  |  | (-0.00461, 0.0424) | 0.115 |
| HDL cholesterol | Binary Exposure | 0.356 | 128,955 | 1,030,579 | (0.332, 0.380) | $9.48 \times 10^{-164}$ |
| | | 0.286 | | | (0.262, 0.310) | $2.40 \times 10^{-108}$ |
| HDL cholesterol | Townsend Index<br>Covariate | 0.350 | 128,955 | 1,030,579 | (0.342, 0.357) | $2.12 \times 10^{-163}$ |
| | | 0.218 | | | (0.212, 0.224) | $1.57 \times 10^{-70}$ |
| Apolipoprotein A | Alcohol Permuted<br>(drinkers only) | 0.312 | 128,161 | 1,030,579 | (0.288, 0.336) | $6.03 \times 10^{-125}$ |
| | | 0.0982 | | | (0.0745, 0.122) | $3.37 \times 10^{-15}$ |
| Apolipoprotein A | Alcohol Permuted | 0.320 | 128,161 | 1,030,579 | (0.296, 0.344) | $2.41 \times 10^{-129}$ |
|  |  | -0.00147 |  |  | (-0.0251, 0.0222) | 0.911 |
| Apolipoprotein A | Random Exposure | 0.312 | 128,161 | 1,030,579 | (0.295, 0.344) | $6.26 \times 10^{-129}$ |
|  |  | -0.00224 |  |  | (-0.0259, 0.0214) | 0.863 |
| Apolipoprotein A | Binary Exposure | 0.308 | 128,161 | 1,030,579 | (0.284, 0.332) | $8.11 \times 10^{-123}$ |
| | | 0.260 | | | (0.236, 0.283) | $3.42 \times 10^{-89}$ |
| Apolipoprotein A | Townsend Index<br>Covariate | 0.302 | 128,161 | 1,030,579 | (0.295, 0.309) | $5.44 \times 10^{-125}$ |
| | | 0.218 | | | (0.212, 0.224) | $1.53 \times 10^{-70}$ |

**Heritability and G×D estimates with varying Alcohol exposure adjustments and SNP selection.** A series of sensitivity tests were applied with varying alcohol exposure adjustments and ran with MonsterLM. Townsend Index covariate refers to the residualization of the Townsend deprivation index as a phenotype covariate.

**Extended Data Table 9.** BMI GWAS Recovery Analyses for BMI, HDL-cholesterol, and Apolipoprotein A with Alcohol.

| GWAS | GWAS SNPs<br>Threshold | Estimate<br>($h_G^2$, G×D) | Individuals, $n$ | SNPs, $m$ | 95% CIs<br>($h_G^2$, G×D) | % Recovered<br>($h_G^2$, G×D) |
| --- | --- | --- | --- | --- | --- | --- |
| Outcome: BMI; G×D: G×Alcohol |  |  |  |  |  |  |
| BMI (GIANT)<sup>31</sup> | $p < 5 \times 10^{-8}$ | 0.0192<br>0.0012 | 141,144 | 374 | (0.018, 0.021)<br>(0.001, 0.002) | 6.13<br>0.70 |
| BMI (GIANT) | $p < 1 \times 10^{-7}$ | 0.0200<br>0.0013 | 141,144 | 407 | (0.019, 0.022)<br>(0.001, 0.002) | 6.38<br>0.80 |
| BMI (GIANT) | $p < 5 \times 10^{-2}$ | 0.1401<br>0.0097 | 141,144 | 45,463 | (0.1345, 0.1457)<br>(0.0052, 0.0143) | 44.62<br>5.86 |
| BMI (GIANT) | $p < 1 \times 10^{-1}$ | 0.1738<br>0.0141 | 141,144 | 80,712 | (0.1666, 0.1809)<br>(0.0081, 0.0202) | 55.34<br>8.51 |
| Alcohol (GSCAN)<sup>29</sup> | $p < 5 \times 10^{-8}$ | 0.00813<br>0.00200 | 141,144 | 731 | (0.007, 0.009)<br>(0.001, 0.003) | 2.59<br>1.21 |
| Alcohol (GSCAN)<sup>29</sup> | $p < 1 \times 10^{-7}$ | 0.00879<br>0.00197 | 141,144 | 840 | (0.008, 0.010)<br>(0.001, 0.003) | 2.80<br>1.19 |
| Alcohol (GSCAN)<sup>30</sup> | $p < 5 \times 10^{-2}$ | 0.185<br>0.024 | 141,144 | 143,078 | (0.177, 0.194)<br>(0.0157, 0.0320) | 59.07<br>14.37 |
| Alcohol (GSCAN)<sup>29</sup> | $p < 1 \times 10^{-1}$ | 0.227<br>0.027 | 141,144 | 228,804 | (0.216, 0.238)<br>(0.0170, 0.0375) | 72.19<br>16.42 |
| HDL-cholesterol<br>(GLGC)<sup>59</sup> | $p < 5 \times 10^{-8}$ | 0.000286<br>0.000863 | 141,144 | 211 | (0.000, 0.001)<br>(0.000, 0.001) | 0.09<br>0.52 |
| HDL-cholesterol<br>(GLGC) | $p < 1 \times 10^{-7}$ | 0.000356<br>0.000902 | 141,144 | 226 | (0.000, 0.007)<br>(0.000, 0.001) | 0.11<br>0.54 |
| HDL-cholesterol<br>(GLGC) | $p < 5 \times 10^{-2}$ | 0.119<br>0.0144 | 141,144 | 71,623 | (0.113, 0.126)<br>(0.008, 0.020) | 38.04<br>8.68 |
| HDL-cholesterol<br>(GLGC) | $p < 1 \times 10^{-1}$ | 0.176<br>0.0210 | 141,144 | 135,470 | (0.167, 0.185)<br>(0.013, 0.029) | 56.07<br>12.63 |
| No GWAS | $p < 1$ | 0.3140<br>0.1663 | 141,144 | 1,030,579 | (0.2920, 0.3359)<br>(0.1446, 0.1879) | 100<br>100 |
| Outcome: HDL-cholesterol; G×D: G×Alcohol |  |  |  |  |  |  |
| Alcohol (GSCAN)<sup>29</sup> | $p < 5 \times 10^{-8}$ | 0.00530<br>0.00178 | 128,955 | 731 | (0.0043, 0.0063)<br>(0.001, 0.002) | 1.51<br>0.63 |
| Alcohol (GSCAN) | $p < 1 \times 10^{-7}$ | 0.00596<br>0.00158 | 128,955 | 840 | (0.005, 0.007)<br>(0.001, 0.002) | 1.70<br>0.75 |
| Alcohol (GSCAN) | $p < 5 \times 10^{-2}$ | 0.179<br>0.0260 | 128,955 | 143,078 | (0.169, 0.188)<br>(0.017, 0.03489) | 51.04<br>12.31 |
| Alcohol (GSCAN) | $p < 1 \times 10^{-1}$ | 0.221<br>0.0453 | 128,955 | 228,804 | (0.209, 0.232)<br>(0.0342, 0.0563) | 63.02<br>21.45 |
| HDL-cholesterol (GLGC)<sup>59</sup> | $p < 5 \times 10^{-8}$ | 0.0984<br>0.00114 | 128,955 | 211 | (0.95, 0.101)<br>(0.000, 0.00162) | 28.11<br>0.54 |
| HDL-cholesterol (GLGC) | $p < 1 \times 10^{-7}$ | 0.0990<br>0.00113 | 128,955 | 226 | (0.0959, 0.102)<br>(0.000, 0.00161) | 28.28<br>0.54 |
| HDL-cholesterol (GLGC) | $p < 5 \times 10^{-2}$ | 0.223<br>0.0153 | 128,955 | 71,623 | (0.215, 0.230)<br>(0.0009, 0.0214) | 63.66<br>7.23 |
| HDL-cholesterol (GLGC) | $p < 1 \times 10^{-1}$ | 0.258<br>0.0315 | 128,955 | 135,470 | (0.249, 0.268)<br>(0.0229, 0.0401) | 73.83<br>14.92 |
| No GWAS | $p < 1$ | 0.350<br>0.212 | 128,955 | 1,030,579 | (0.188, 0.235)<br>(0.326, 0.373) | 100<br>100 |
| Outcome: Apolipoprotein A; G×D: G×Alcohol |  |  |  |  |  |  |
| Alcohol (GSCAN)<sup>29</sup> | $p < 5 \times 10^{-8}$ | 0.00423<br>0.00157 | 128,161 | 731 | (0.003, 0.005)<br>(0.001, 0.002) | 1.40<br>0.74 |
| Alcohol (GSCAN) | $p < 1 \times 10^{-7}$ | 0.00520<br>0.00150 | 128,161 | 840 | (0.004, 0.006)<br>(0.001, 0.002) | 1.72<br>0.70 |
| Alcohol (GSCAN) | $p < 5 \times 10^{-2}$ | 0.121<br>0.0299 | 128,161 | 143,078 | (0.114, 0.129)<br>(0.0204, 0.0394) | 40.07<br>14.04 |
| Alcohol (GSCAN) | $p < 1 \times 10^{-1}$ | 0.193<br>0.0445 | 128,161 | 228,804 | (0.182, 0.205)<br>(0.0333, 0.0556) | 63.98<br>20.87 |
| HDL-cholesterol (GLGC)<sup>59</sup> | $p < 5 \times 10^{-8}$ | 0.0799<br>0.000877 | 128,161 | 211 | (0.0771, 0.0828)<br>(0.000, 0.00132) | 26.47<br>0.41 |
| HDL-cholesterol (GLGC) | $p < 1 \times 10^{-7}$ | 0.0807<br>0.000858 | 128,161 | 226 | (0.0779, 0.0836)<br>(0.000, 0.001) | 26.73<br>0.40 |
| HDL-cholesterol (GLGC) | $p < 5 \times 10^{-2}$ | 0.191<br>0.0161 | 128,161 | 71,623 | (0.183, 0.198)<br>(0.010, 0.022) | 63.14<br>7.58 |
| HDL-cholesterol (GLGC) | $p < 1 \times 10^{-1}$ | 0.225<br>0.0303 | 128,161 | 135,470 | (0.215, 0.234)<br>(0.022, 0.039) | 74.36<br>14.24 |
| No GWAS | $p < 1$ | 0.302<br>0.213 | 128,161 | 1,030,579 | (0.190, 0.237)<br>(0.279, 0.326) | 100<br>100 |

**Heritability and G×D estimates with varying SNP selections.** SNPs were selected based off their respective GWAS study association levels. MonsterLM was used to compute heritability and G×D with the SNPs from each threshold.

**Extended Data Table 10.** Key Definitions for the MonsterLM G×D Framework.

| Term | Definition |
| --- | --- |
| Genome-wide G×D | The variance in an outcome explained by genome-wide genotype-by-diet interactions, estimated as adjusted R<sup>2</sup> across all SNPs using the MonsterLM framework. |
| G×D interaction weights ($\beta_{G \times D}$) | SNP-specific interaction effect estimates derived from the discovery set, representing the regression coefficient for the genotype-by-diet interaction term. |
| G×D polygenic score (G×D score) | A score constructed by summing SNP genotypes weighted by their corresponding interaction effect estimates derived from the discovery set and applied in the validation set. |
| Unrealized G×D score | A G×D score calculated without incorporating the dietary exposure, representing the sum of interaction-weighted genetic variants for each individual. |
| Realized G×D score | A G×D score calculated by multiplying the interaction-weighted genetic variants by the corresponding dietary exposure for each individual, thereby incorporating the gene–diet interaction term. |
| Interaction-based score (vs traditional PRS) | A polygenic score based on interaction effect estimates rather than marginal genetic effects, capturing gene–diet interaction effects rather than direct genetic associations with the outcome. |

Definitions for terminology in the MonsterLM G×D analytical framework.

